# A rare genetic disorder provides insights into mechanisms of early-onset neurodegeneration

**DOI:** 10.1101/2023.05.22.23290124

**Authors:** Cherif Badja, Sophie Momen, Gene Ching Cheik Koh, Soraya Boushaki, Theodoros I. Roumeliotis, Zuza Kozik, Ian Jones, Vicky Bousgouni, João M. L. Dias, Marios G. Krokidis, Jamie Young, Hongwei Chen, Ming Yang, France Docquier, Yasin Memari, Lorea Valcarcel-Jimenez, Komal Gupta, Li Ren Kong, Heather Fawcett, Florian Robert, Salome Zhao, Andrea Degasperi, Helen Davies, Rebecca Harris, Christian Frezza, Chryssostomos Chatgilialoglu, Robert Sarkany, Alan Lehmann, Chris Bakal, Jyoti Choudhary, Hiva Fassihi, Serena Nik-Zainal

**Author notes:** These authors contributed equally (Cherif Badja, Sophie Momen).

## Abstract

Xeroderma pigmentosum (XP) is characterized by defective repair of ultraviolet radiation(UVR)-induced DNA damage. Patients have UVR hypersensitivity and increased skin cancer risk. Effective photoprotection has reduced childhood cancer-related deaths, but revealed adolescence-onset neurodegeneration, arising through unknown mechanisms. Here, we investigate XP neurodegeneration using pluripotent stem cells derived from XP patients and healthy relatives, performing functional multi-omics on samples during neuronal differentiation. We find endoplasmic reticulum stress is upregulated, preceded by oxidative stress, causing substantial 5’,8-cyclopurine and 8-oxopurine DNA damage. Critically, XP neurons exhibit inappropriate downregulation of the protein clearance ubiquitin-proteasome system (UPS). Chemical enhancement of UPS activity improves phenotypes, albeit inadequately, implying that early detection/prevention strategies are necessary to produce clinically impactful outcomes. Thus, we develop an early detection assay predicting neurodegeneration in at-risk patients.

## Main text

Patients with the genetic condition xeroderma pigmentosum (XP) are deficient in nucleotide excision repair (NER)^1, 2^. NER plays a fundamental role in repairing bulky DNA damage caused by ultraviolet radiation (UVR)^3–5^. XP patients thus have reduced tolerance to UVR, exhibiting exaggerated sunburn in ∼50% of cases^6, 7^, pigmentary changes, ocular damage, and carry a 10,000-fold increased risk of cutaneous malignancies^6, 8^. Mutations in multiple genes, *XPA*, *XPB/ERCC3*, *XPC*, *XPD/ERCC2*, *XPE/DDB2*, *XPF/ERCC* and *XPG/ERCC5* lead to defective NER, while mutations in *XPV* manifest a later-onset clinical course^9^.

Early biological understanding of skin and eye features of XP prompted preventative clinical interventions including photo-protection, dermatological and ophthalmological surveillance, and treatment of premalignant lesions^10^. While these measures considerably reduced premature mortality from metastatic skin cancers^11^, approximately one third of XP patients subsequently develop progressive neurodegenerative symptoms in adolescence and early adulthood, including sensorineural hearing loss, axonal neuropathies, hyporeflexia, and cognitive impairment, affecting quality of life and survival^6, 12^. Today, the mechanistic basis of XP neurological deterioration remains poorly understood, with no effective means of preventing or treating this devastating, fatal XP complication.

Since exogenous UVR cannot reach the human brain, endogenous bulky DNA lesions called 5’,8-cyclopurine-2’-deoxynucleosides (herewith, cPus) have been hypothesized to elicit XP related neurodegeneration^13–16^. These oxidative lesions are repaired exclusively by NER; how they precipitate neuronal dysfunction, however, is unclear.

Here, we investigate patients from multiple XP complementation groups using human induced pluripotent stem cell (hiPSC) models. We demonstrate how systematic, functional multiomic characterization at several stages of neuronal differentiation can reveal striking, coordinated, homeostatic pathway abnormalities that are powerfully consistent across replicates of XP genotypes. We show accumulation of oxidized nucleosides in XP neurons for the first time. Intriguingly, the ubiquitin proteasome system is inappropriately downregulated in early-onset XP neurodegeneration, presenting new insights with intervention potential.

### Endoplasmic Reticulum stress/Unfolded Protein Response (ER stress/UPR) is upregulated in XP neuronal models

We derived hiPSC models from four XP patients with neurodegeneration; one male XPD patient (NXPD-30), two male and one female XPG patients (NXPG-34, NXPG-43, NXPG-32), and a healthy control (CTRL-33, unaffected heterozygous relative of NXPG-32 and NXPG-34). We performed directed differentiation into neurons in triplicate (R1, R2 and R3) for each genotype (Fig. 1a). We collected cell pellets at multiple differentiation stages: hiPSC day 0 (d0), precursor neuronal stem cells (NSCs) day 6 (d6) and day 12 (d12), and terminally differentiated neurons at day 27 (d27) and day 42 (d42) respectively (Fig. 1a, Fig. S1a-e).

**Fig. 1.**
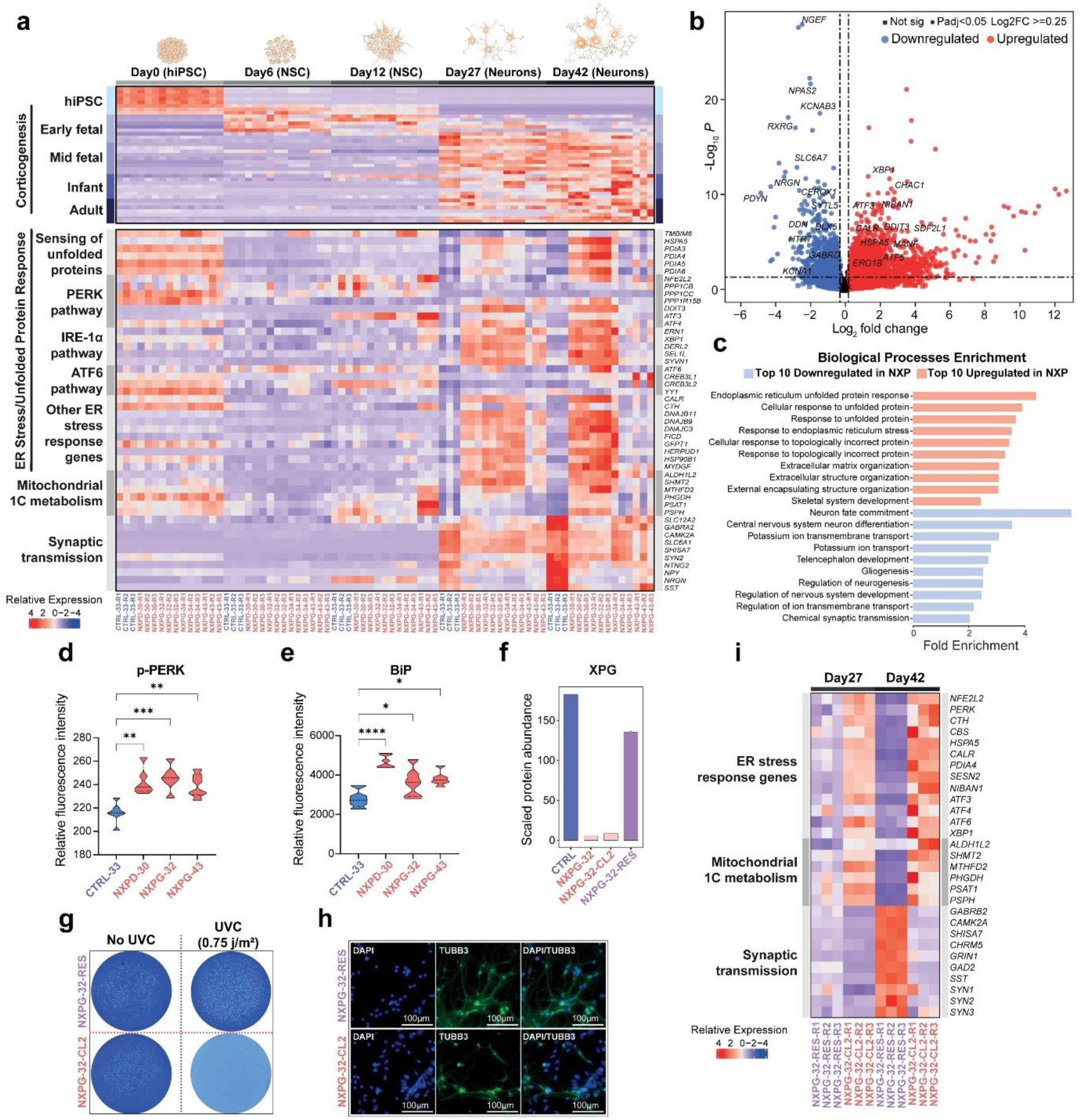
ER stress/UPR is upregulated in neuronal models of XP and genetic correction reverts the phenotype. **(a)** Heatmap of relative transcriptomic levels at the pluripotent stem cell stage (Day 0) through directed differentiation (Day 6 and Day 12, NSC stage) into neurons (Day 27 and Day 42), from left to right. Upper panel shows genes implicated in pluripotency and various stages of human corticogenesis. Lower panel shows genes and pathways involved in proteostasis (ER stress/UPR). X-axis: healthy, heterozygous control in blue (CTRL-33-xx); XP patients with neurodegeneration in red (NXPx-xx-xx). Three replicates available for each genotype (R1, R2 and R3). **(b)** Volcano plot of DEG analysis of Day 42 neurons from XP patients with neurodegeneration (NXP) versus control. **(c)** Enrichment of biological processes based on DEGs contrasting Day 42 NXP neurons to control. **(d)** Quantitative immunofluorescence (QIF) measurements of phospho-PERK (p-PERK) performed on neurons derived from NXP patients (red) and control (blue) (One-way ANOVA, **: p<0.01, ***: p<0.001). **(e)** QIF of BiP performed on NXP neurons (red) and control (blue) (One-way ANOVA, *: p<0.05, ****: p<0.0001). **(f)** Scaled protein abundance of XPG in control (blue), XP case with neurodegeneration (NXPG-32), XP case with CRISPR-Cas12-corrected alleles (NXPG-32-RES) and a clone that has been through the CRISPR-Cas12 process but has not had a corrected allele (i.e., full biallelic mutant, NXPG-32-CL2). **(g)** Colony forming assay contrasting responses of *XPG* mutant hiPSC (NXPG-32-CL2) to rescued hiPSC (NXPG-32-RES) with and without UVC irradiation (0.75j/m^2^). **(h)** Immunofluorescence characterisation of rescued cells (NXPG-32-RES) and uncorrected *XPG* mutant cells (NXPG-32-CL2) using DAPI and TUBB3 (neuronal marker). **(i)** Transcriptomic heatmap of genes involved in proteostasis, mitochondrial 1C metabolism and synaptic transmission demonstrating phenotypic recovery in rescued cells.

First, we explored bulk transcriptomics contrasting d42 neurons from XP patients (herewith, XP neurons) to controls (Fig. 1b). 3,351 differentially expressed genes (DEGs) (padj <0.05) were identified with 2,044 upregulated and 1,307 downregulated. Gene Ontology (GO) annotation of DEGs revealed upregulation of the endoplasmic reticulum (ER) stress response, and downregulation of neuronal maturation and synaptic signaling (Fig. 1c).

The ER is a major protein folding compartment critical to protein homeostasis (or proteostasis)^17^. Imbalances between protein-folding demand and folding capacity results in accumulation of misfolded/unfolded proteins or ER stress, triggering the Unfolded Protein Response (UPR)^18^, an adaptive signaling pathway that stimulates protein clearance^19, 20^ via three transmembrane signaling proteins; PERK^20^, ATF6^21^, and IRE-1a^22^, which are usually bound to BiP^23^. During ER stress, BiP is titrated away inducing protein clearance^23^. Loss of proteostasis is implicated in cancer and neurodegeneration^24–28^, and may be relevant in XP neurodegeneration.

We extended bulk transcriptomics to all stages of differentiation (Fig. 1a, Fig. S1a; Table S1). UPR activity was not distinguishing between XP patients and controls (Fig. 1a) at pluripotency (d0) and NSC stages (d6 and d12) (Fig. S1b). However, upon terminal differentiation (d27 and d42), quantitative gene expression of all three UPR branches showed striking, progressive upregulation in all technical replicates of all four XP patients (Figure 1a, lower panel), in contrast to markers of generic differentiation which were similar between XP patients and controls (Fig. 1a, top panel).

For validation, we performed quantitative immunofluorescence (QIF) observing significant elevation of BiP (padj<0.045) and PERK/phosphorylated PERK (padj<0.0047) in XP neurons when contrasted to controls (Fig. 1d, 1e, Fig. S1f-i). Furthermore, our models showed upregulated serine biosynthesis pathway enzymes (*PHGDH*, *PSAT1* and *PSPH*) and associated elevation of mitochondrial 1C metabolism (*SHMT2*, *MTFHD2* and *ALDH1L2*) in line with previous work, where ER stress was artificially incited^29^.

Finally, we restored mutant XPG alleles to wildtype in NXPG-32 hiPSCs using an efficient CRISPR-Cas12 strategy. Cells with restored alleles (NXPG-32-RES) were contrasted to NXPG-32-CL2 cells that have been through the CRISPR strategy but not rescued. Genotypes were confirmed by Sanger sequencing (Fig. S2a), functional verification obtained via proteomics (Fig. 1f), Western blotting (Fig. S2b), and colony formation assays post-UVC irradiation (Fig. 2g, Fig. S2c). Taking both rescued and uncorrected cells in triplicate through neuronal differentiation (Fig. 1h), we found that rescued cells were indistinguishable from wildtype, morphologically and transcriptionally, in pluripotent and differentiated neuronal states (Fig. 1i, Fig. S2d-f, Table S2). Rescued NXPG-32-RES cells had normal proteostasis in contrast to mutant NXPG-32-CL2, confirming that ER stress/UPR upregulation in XP neuronal models is directly linked to the underlying NER genetic abnormality (Table S2).

**Fig. 2.**
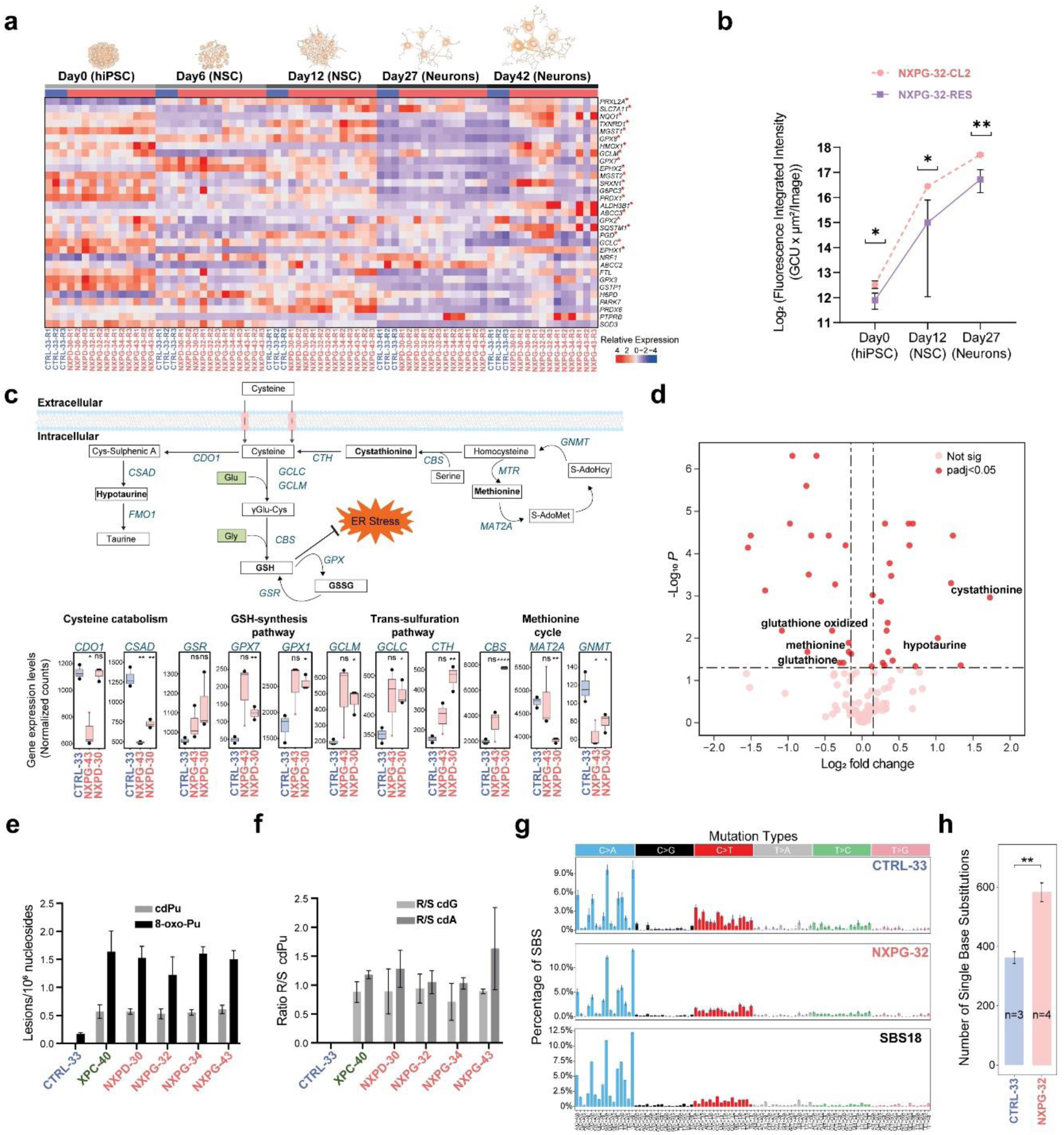
Oxidative stress precedes ER stress/UPR, is sustained through directed differentiation, and causes substantial DNA damage. **(a)** Transcriptomic heatmap of downstream targets of NRF2 showing upregulation at earlier differentiation timepoints, including at NSC and hiPSC stages. Significantly differentially expressed NRF2 targets are highlighted with “*”. **(b)** ROS assay on NXP (NXPG-32-CL2) cells and a CRISPR-corrected clone (NXPG-32-RES) (p< 0.03, unpaired t-test). **(c)** Schematic of metabolomic pathways implicated in the anti-oxidant response. Shift in various metabolites and enzymes from trans-sulfuration to GSH-synthesis pathway given chronic oxidative stress (p<0.01*, p<0.01**, t-test). **(d)** Volcano plot of metabolomics of NXPD-30 and NXPG-42 compared to CTRL-33 Day 42 neuronal replicates highlighting critical metabolomic factors that are distinguishing. **(e)** Levels of 5’,8-cyclopurine (cdPu) and 8-oxopurine (8-oxo-Pu) lesions in XP samples with neurodegeneration (red), XP without neurodegeneration (green) and controls (blue) (lesions/10^6^ nucleosides). Values are mean ± SD (n=3). **(f)** Diastereoisomeric ratios (5’*R*/5’*S*) for cdG and cdA lesions in XP samples with neurodegeneration (red), XP without neurodegeneration (green) and controls (blue). The numbers represent the mean value (± standard deviation) in triplicate. **(g)** Substitution mutations in WGS clones of control (CTRL-33) and XP patient (NXPG-32). Comparison to SBS18 previously reported in failure of repair by *OGG1*. Cosine similarity between WGS clones to SBS18 was 0.94 and 0.96, respectively, suggesting that mutagenesis is driven qualitatively by oxidative species in control and XP cases. **(h)** Counts of substitution mutations showing quantitative differences between XP cases and controls. (t-test, p<0.01**).

### Oxidative stress precedes ER stress/UPR in XP neurons

UPR can be provoked by oxidative stress^30^, a postulated source of DNA damage in XP-related neurodegeneration^14–16, 31^. If oxidative stress triggers UPR, it would manifest temporally before ER stress. Interestingly, Nuclear Factor Erythroid 2-related factor 1-2 (*NFE2L2,* aka *NRF2),* a major antioxidant response regulator^32^, is upregulated earlier by d12 (NSC stage) preceding ER stress/UPR (Fig. 1a). Furthermore, downstream targets of *NFE2L2* are universally upregulated at d12 (Fig. 2a) and remain upregulated at d42. To confirm oxidative stress, we measured cellular ROS using fluorescence microscopy (Fig. 2b). Background levels of ROS were elevated in hiPSCs, intensified at d12 (NSC) and remained elevated in d27 neurons of rescued NXPG-32-RES and mutant NXPG-32-CL2 cells, although the latter were significantly more adversely affected (p<0.03). Oxidative stress therefore preceded ER stress, was sustained throughout differentiation, and the mounting ER stress/UPR may be an adaptive response to the interminable oxidative assault.

To seek corroborating evidence of adaptation to chronic oxidative stress, we examined the metabolic state of d42 neurons from two XP patients (*XPD* and *XPG* mutants, five replicates each) contrasting them to controls. Metabolomics integrated with gene expression data suggested a strong shift from trans-sulfuration towards synthesis of glutathione, a potent mammalian antioxidant (Fig. 2c), evidenced by marked upregulation of cystathionine, a component of glutathione, and associated reduction of methionine (Fig. 2d, Fig. S3a). In addition, cystathionine beta-synthase (CBS) and cystathionine gamma-lyase (CTH) required for multi-step generation of glutathione, were both elevated, driving the trans-sulfuration shift towards glutathione synthesis. Glutathione exists in the reduced thiol state (GSH) and is oxidized into glutathione disulphide (GSSG) during oxidative stress. Despite considerable synthesis, GSH levels remained depleted (reflected in GSSG/GSH ratios that are non significant), implying that chronic, overwhelming oxidative stress was exhausting the GSH pool in XP neurons, in spite of increased GSH generation.

### Oxidative stress causes purine DNA damage in XP neurons

Oxidative stress triggers cPu formation, the postulated culprit inducing neurological XP^14, 15^. cPus form when hydroxyl radicals react with 2’-deoxy-guanosine (2’-dG) or 2’-deoxy adenosine (2’-dA)^14, 33^ (Fig. S3b) causing structural deformation, creating a bulky substrate for NER^34^. Two diastereomers can be produced, 5’*R* and 5’*S*, depending on H and OH stereochemistry at the chiral 5’ carbon^15, 33, 35^ (Fig. S3b). 5’*R* diastereomers are more efficiently repaired by NER, with ratios of 5’*S*:5’*R* ∼2:1 in human cell-free extracts^36^.

To quantify oxidized nucleosides, we performed LC-ESI-MS/MS (liquid chromatography electrospray ionisation tandem mass spectrometry) on neurons from XP patients with (NXP) and without neurodegeneration (XP-non), and healthy controls. Remarkably, we found substantially elevated cPus in XP neurons (irrespective of whether NXP or XP-non) compared with controls (Fig. 2e, p<0.002). Additionally, the 5’*S*:5’*R* diastereomer ratio in XP neurons was ∼1:1 (Fig. 2f), concordant with deficient NER repair of 5’*R*, resulting in 5’*R* accumulation and near equivalence with 5’*S* diastereomers.

Surprisingly, we also found increased 8-oxopurines in XP neurons (Fig. 2e, Fig. S3c). Although absolute 8-oxopurine values exceeded cPus, the fold increase was greater for cPus than 8-oxopurines, resulting in an observed cPu:8-oxopurine ratio of 1:2-1:3 rather than the expected 1:40^37, 38^, similar to previously reported levels in XPA-deficient epithelial embryonic cells^39^. Note that 8-oxopurines are observed in controls, in-keeping with prior reports that hiPSCs are exposed to oxidative damage^40–42^.

These results support the hypothesis that cPus contribute to DNA damage in NER-deficient neurons. The increased 8-oxopurines is intriguing, given that these lesions are BER repaired^43^. Perhaps NER plays a greater role in 8-oxopurine repair than previously appreciated. An alternative explanation is that such marked oxidative stress induces multiple types of oxidative lesions^44^ including 8-oxopurines, with a greater relative impact on cPus in XP, because of the exclusive role played by NER in sanitizing cPus.

Lastly, to demonstrate that oxidative stress could impart lasting DNA damage and mutations, we performed whole genome sequencing of single-cell subclones derived from NSCs of NXPG-32 and CTRL-33 (Fig. 2g-h). Both sets of NSCs showed substitution Signature 18 mutations, associated with failure of clearing 8-oxo-dG lesions^45^. XP patients showed a significantly higher burden of signature 18 (p<0.01, t-test), reinforcing the observation that XP cells suffer increased oxidative stress.

### Ubiquitin-proteasomal dysfunction is implicated in XP neurodegeneration

Although DNA oxidative lesions accumulate in neurons of all XP patients, only some XP patients develop neurological disease. To explore post-translational pathophysiology discriminating NXP from XP-non, we generated proteomics on d27 neurons (Fig. S4a-c). From 8,352 quantified proteins, we first contrasted four NXP to four controls, identifying 468 differentially expressed proteins (DEPs, p<0.05) (Fig. 3a, Table S3). We also contrasted a larger cohort of eight NXP to eight XP-non cases and found 949 DEPs (p<0.05) (Fig. 3b, Table S3). Pathway enrichment analyses of both datasets revealed consistent differentially enriched biological processes: upregulation of ubiquitin-dependent protein catabolic processes, the Golgi transport complex and mitotic cell cycle, all anticipated in the context of increased ER stress; downregulation of synaptic/neurotransmitter activity and axon guidance, probable consequences of neuronal dysfunction (also validating transcriptomic results); but notably, downregulation of the ubiquitin proteasome system (UPS), an unexpected finding (Fig. 3c).

**Fig. 3.**
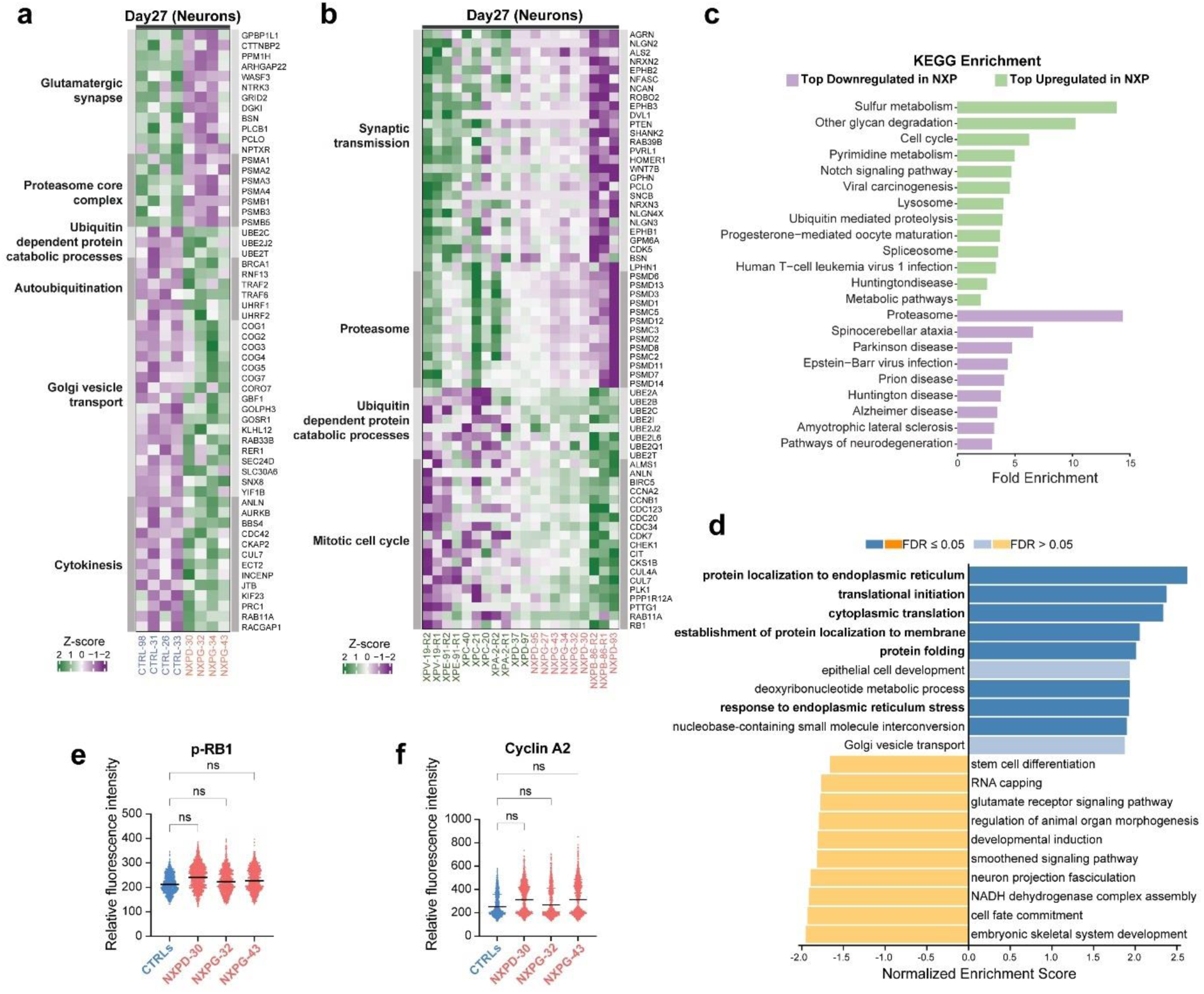
Ubiquitin-proteasomal system (UPS) dysregulation is implicated in XP neurodegeneration. **(a)** Proteomic data comparing neurodegenerative XP cases against control highlighting representative proteins from pathways that have the most significantly differentially expressed proteins. **(b)** Proteomics comparing neurodegenerative XP cases (NXP, red) against XP without neurodegeneration (XP-non, green). **(c)** KEGG enrichment demonstrating the ten most distinguishing upregulated and downregulated pathways when contrasting NXP to XP-non. **(d)** Themes enriched in strongly positively (blue, top) and negatively (yellow, bottom) proteins that correlate with QIF p-PERK expression as determined through GSEA (P< 0.05, FDR labelled). **(e)** Relative fluorescence intensity of QIF of phosphoRB1 (p-RB1) in control and XP cells. **(f)** Relative fluorescence intensity of QIF of Cyclin A2 in control and XP cells (ns= not statistically significant).

Normally, the ER eliminates misfolded/unfolded proteins by translocation to the cytoplasm, targeting them for polyubiquitination and UPS degradation^46^. This initiates protein clearance including triggering markers of cell cycle and de-differentiation^47^. To seek support for whether these downstream proteomic observations were related to ER stress, we sought the relationship between QIF of phosphorylated PERK (p-PERK) and the proteome using partial-least squares regression analyses. We found that higher p-PERK concentrations (indicative of ER stress) were associated with deranged proteostasis: ‘protein localisation to endoplasmic reticulum’, ‘translational initiation’, ‘cytoplasmic translation’ ‘protein folding’, (FDR < 0.05, including BiP, CALR, CANX) (Fig. 3d), and reduced expression pertaining to ubiquitin-related protein catabolism (biological process; ‘ubiquitin-dependent protein catabolic process’, ‘protein catabolic process’, (FDR< 0.05), including PSMB1, UBA6, UBE2E1/UBE2E2, CUL2/CUL9) (Fig. S4d). Over-enrichment analysis also implicated ‘cell cycle processes’ (FDR< 0.05, including; TOP2A, CCNB1, ECT2) (Fig. S4e). These integrated analyses were consistent with a direct relationship between QIF ER stress and proteomic findings.

To confirm that observed upregulation of cell cycle markers are consequences of ER stress and not indicative of mitotic re-entry, we used high content image analysis measuring two cell cycle proteins (Fig. 3e,3f). Both phosphoRB1 (p-RB1), signifying G1 transition, and cyclin A2, denoting S phase transition, showed no difference in levels between controls and patients.

The most striking finding was UPS downregulation. The 26S proteasome comprises a hollow 20S core particle housing a protein degradation catalytic cavity and two 19S regulatory cap subunits, which have ATPase activity and polyubiquitinated protein recognition sites (Fig. 4a)^48^. We examined 26S proteasome component abundances. We found significant downregulation of 19S lid and base regulatory subunits in NXP versus XP-non. In addition, PSMF1, a proteasome inhibitor subunit is upregulated in NXP (Fig. 4b). The 20S core, activator subunits, and assembling chaperone proteins were not distinguishing.

**Fig. 4.**
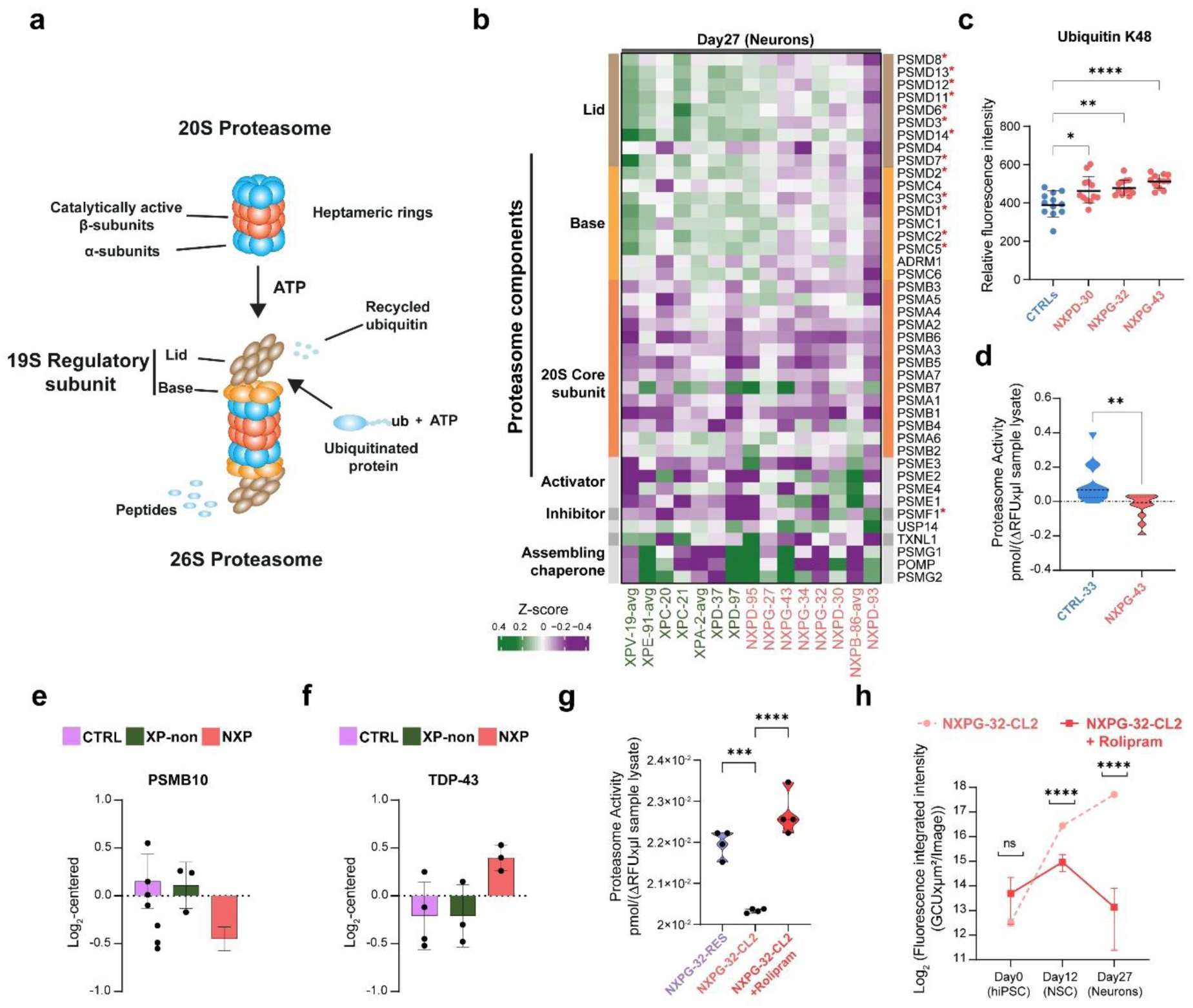
26S proteasome capacity is compromised, can be enhanced through small molecule therapy but is inadequate to salvage the neuronal phenotype. **(a)** Schematic representation of the proteasome. **(b)** Protein expression of components of the proteasome (“*” are significant DEPs when NXP and XP-non are contrasted). **(c)** Polyubiquitination of K48 in several XP cases and several controls (One-way ANOVA, *: p<0.02, **: p<0.002, ****: p<0.0001). **(d)** Proteasomal activity assay in neurons at day 27 contrasting an XP case against control (One-way ANOVA, **: padj <0.007). **(e)** Immunoprecipitation (IP) followed by proteomics quantification of PSMA1 interactors showing decreased association of PSMB10 with the proteasome in NXP (n=3) neurons at day 27 compared to controls (n=4) and XP-non (n=3). (Student’s t-test, p<0.05)**. (f)** IP followed by proteomics quantification of PSMA1 interactors showing increased association of TDP-43 with the proteasome in NXP (n=3) neurons at day27 compared to controls (n=4) and XP-non (n=3). (Student’s t-test, p<0.05). **(g)** Proteasomal activity assay in neurons at day 27 contrasting an XP case against her CRISPR Cas12 corrected model (One-way ANOVA, *: padj <0.05, ***: padj <0.0005, ****: padj <0.0001). **(h)** ROS assay on NXPG-32-CL2 cells untreated and treated with 30 µM Rolipram (One way ANOVA, ns: not statistically significant, ****: padj <0.0001).

The decrease in 19S abundance and upregulation of PSMF1 implicate accumulation of proteins tagged for 26S proteasomal degradation. Using ubiquitin K48 linkage antibody (Fig. 4c), we found K48 polyubiquitination excess in NXP neurons compared to controls (padj<0.01), confirming a defect in removal of ubiquitinated substrates. We ratified this with a proteasome fluorescence activity assay corroborating a reduction in proteasomal function in NXP compared to control (Fig. 4d). Also, there was neither transcriptional nor proteomic evidence of compensation by the alternative protein clearance autophagy-lysosomal pathway.

To understand how XP deficiency could contribute to a defect in protein clearance, we performed immunoprecipitation-Mass Spectrometry (IP-MS) using an antibody to PSMA1 (a core component of 20S proteasome) on three NXP patients cell lines, three XP-non and four controls. We recovered all core components of the 20S, as well as components of mature 19S, PA200, PA28a/b and PA28g (Table S4) regulatory subunits, and all three subunits of the immunoproteasome^49^. Intriguingly, PSMB10, a beta-2i catalytic subunit of the 20S proteasome is reduced in NXP (Fig. 4e), even though it is better known for its neuro-inflammation role ^50^. Other neuro-inflammatory components and markers of interferon-signaling were not elevated in NXP suggesting that beyond the immuno-proteome, PSMB10 may play a distinct role in NXP disease progression. Additionally, there was an association between the proteasome and TDP-43 (Fig. 4f), implicated in various neurological diseases^51^. TDP-43 forms aggregates that trap and stall proteasomes in neurons^52^, yet post-mortem brain histology of NXP cases has not revealed gross protein aggregates^53^. Lastly, transcriptomic levels of UPS were not distinguishing between NXP and controls, even though the proteome showed marked differences, underscoring how UPS dysfunction is likely a post-transcriptional phenomenon in NXP.

### Proteasomal activity enhancement in XP neurons brings limited benefit

Given that proteasomal dysfunction distinguishes NXP from XP-non, we asked whether proteasome activity enhancement could be beneficial for NXP patients. We exposed XPG patient cells to 30µM rolipram, a selective phosphodiesterase-4 inhibitor (PDE4i) used in autoimmune and neurodegenerative diseases^54, 55^ at hiPSC stage (day 0). Rolipram treatment reverted proteasome functional activity to levels observed in healthy cells (Fig. 4g). Rolipram treated cells also showed significant relative reduction in ROS levels compared to untreated cells (padj<0.0001, Fig. 4h). However, we note that ROS production was strongly exacerbated by differentiation and absolute ROS levels remained high even with treatment, suggesting that while improvement can be achieved, much of the damage is happening or done, and likely inadequate to reverse a degenerating phenotype.

### Early-detection assay for predicting neurodegeneration in XP

Progressive neurodegeneration affects only some XP patients and is more prevalent amongst *XPA*, *XPB, XPD, XPF* and *XPG* genotypes^6, 12^. We investigated developing a neurodegeneration prediction model using *in vitro* neuronal proteome data to aid clinical management and prognostication.

We used support vector machine-learning (SVM) models to classify neurological statuses of XP patients based on normalized proteomics from seventeen neuronal samples derived from thirteen XP patients (nine single samples, four with replicates). Eight patients were NXP and five were XP-non. A leave-one-out cross-validation approach was used to optimize the model because of the (rarity of the disease and thus) small dataset (Fig. 5a; Table S5). We explored model performance using proteins from the ten most distinguishing biological pathways (154 DEPs) (Table S5). Then, we assessed model accuracy using up to the top 16 most discriminating ranked proteins (Fig. 5a). The highest average accuracy was 88% at distinguishing NXP from XP-non correctly, achieved with nine proteins (Fig. 5b), four with negative SV coefficients (CDC20, ANLN, CEP290 and BBS4) and five with positive coefficients (JADE1, LGMN, INS, BDNF and PPID) (Fig. 5c).

**Fig. 5.**
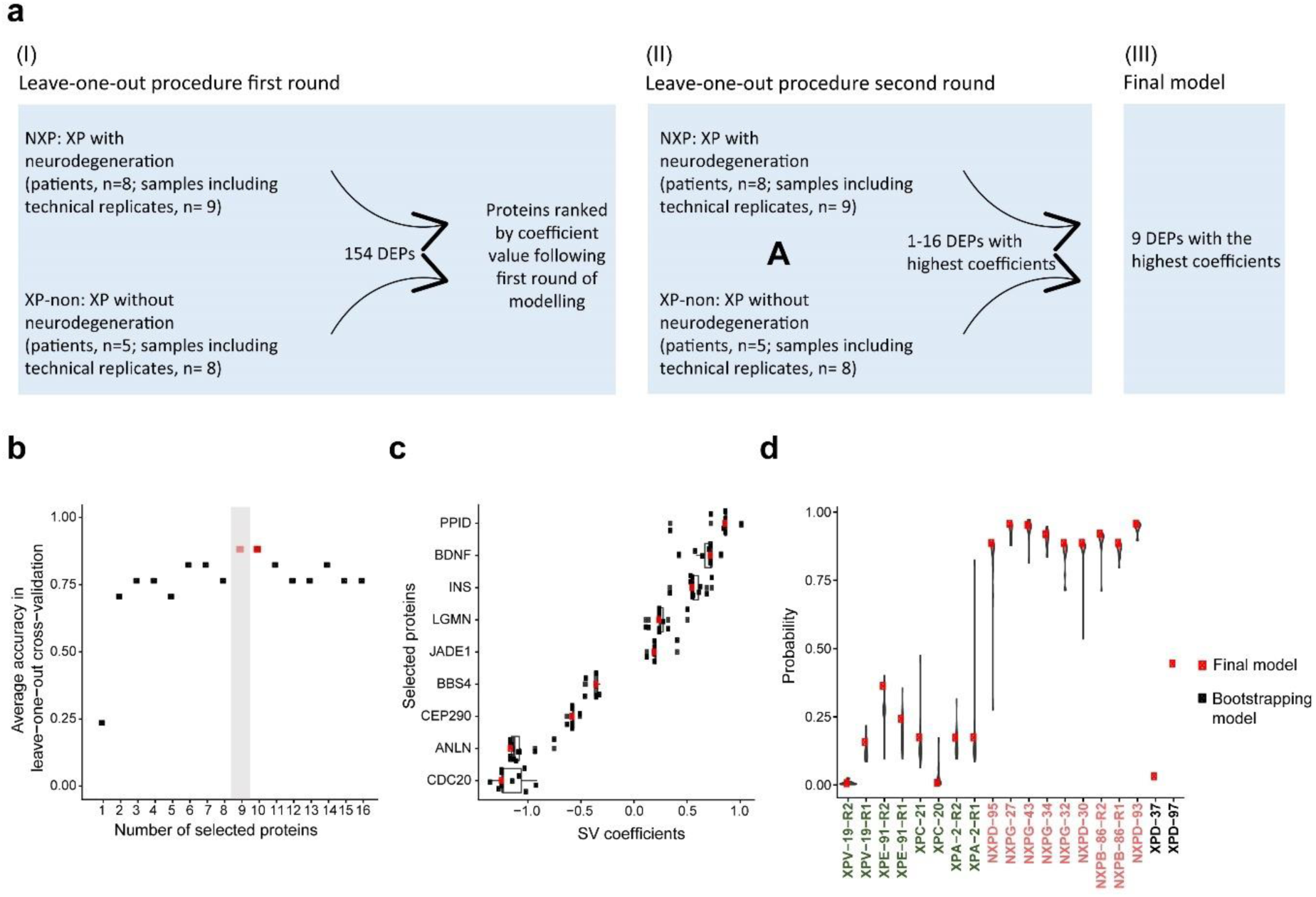
A proteomics-based early detection assay for neurodegeneration in XP. **(a)** Workflow of developing a proteomics based predictive assay using support vector models (SVM) machine learning in a small cohort using a leave-one-out strategy. **(b)** Assessing the accuracy of the model. **(c)** Model predictions based on nine proteins, including the 17 cases used for training and two new cases that are not (so far) manifesting a neurological phenotype. **(d)** SV weights for the nine proteins.

We applied our trained model on two cases (XPD-37 and XPD-97) excluded from the training process because they had not manifested neurological features (so far) but were ‘at risk’ given their *XPD*-mutated genotypes (Fig. 5d). According to our SVM model, XPD-37 was not predicted to develop neurodegeneration. By contrast, XPD-97 was.

In the UK, most XPD patients carry Arg683 mutations, where hydrophilic arginine is replaced by hydrophobic tryptophan (p.Arg683Trp) causing considerable XPD disruption and a severe phenotype^12^. Patient XPD-37 has an arginine to glutamine change, and no neurological abnormalities at age 37. The lack of neurological symptoms in-keeping with the SVM prediction may be due to glutamine being hydrophilic (like arginine). XPD-97 has three inherited mutations. He is heterozygous for the common p.Arg683Trp allele and carries two mutations on the other parental allele, p.Leu461Val and p.Ala717Gly. The latter mutation is predicted to generate a new splice donor site, resulting in possible read-through^56^ with uncertainty regarding phenotype. It is possible that the model correctly predicts XPD-97 to develop neurodegeneration; however, the patient is very young and neurology may not have manifested yet.

Our model could potentially be utilized as a functional assay for predicting progressive neurology, highlighting individuals for early intervention, particularly where a previously unreported XP mutation is identified, and/or where a patient’s clinical picture is out-of-keeping because unknown modifiers may be involved. A longitudinal study (beyond this work) would be required to validate the predictive power of the assay.

## Discussion

Our parallel time-course strategy of independent replicates, contrasting multiple diverse XP genotypes and controls, requires fastidious quality control. Yet coupled to functional multiomic characterization, it is a powerful approach that shows consistent pathway disparities, is quantitative, and can provide dynamic information, including whether certain types of homeostatic dysregulation could temporally precede others.

This work sheds light on a distressing phenomenon of adolescent-onset XP neurodegeneration (Fig. S5). While both ER stress and UPS dysfunction have been implicated in older-onset neurodegenerative disorders, here we show that ER stress/UPR in XP neurons does not manifest at earlier differentiation timepoints, is heralded by oxidative stress, that remains sustained through differentiation. There is chronic maladaptation of neuronal metabolism to continuous oxidative stress, and direct DNA damage with accumulation of 5’,8-cyclopurines and 8-oxopurines in XP neurons. irrespective of whether NXP or XP-non. While this may seem counter-intuitive, DNA damage will accrue regardless of genotype. From a repair perspective, XP-non cases (although defective of global genome NER) have functional transcription coupled NER (TC-NER) and will be able to remove oxidative damage from transcribed genes, a smaller (but critical) subset of the genome, that may be too small a difference to detect with LC-ESI-MS/MS assays. Regardless, the interrelationship of loss of proteostasis, oxidative stress, and oxidative DNA damage are unifying as a cause for accelerated neuronal ageing in XP^28, 57, 58^.

Critically, in patients with neurodegeneration, one pathway fails to respond to upstream pathophysiology: the UPS. UPS dysregulation in XP is post-transcriptional; 26S proteasomal presence is globally reduced and the PSMB10 subunit, previously associated with the immuno proteome is specifically affected. IP has not revealed direct interactions between the proteasome and XP proteins, thus further work is required to understand how NER deficiency leads to selective UPS dysfunction.

Small molecule inhibition to enhance proteasome activity substantiates the role played by dysfunctional UPS in XP neurodegeneration. Per contra, the level of reduction in ROS production was significant, but not enough and likely too late to prevent the damaging phenotype. This raises an important clinical argument treating a downstream pathophysiological target once a patient has manifested neurodegenerative symptoms is a belated, flawed approach. By that point, disease pathology is considerable at the cellular level. A more valuable approach is to identify at-risk individuals early, prior to neurodegeneration, and intervene with preventative measures from early life.

Indeed, our functional assay potentially offers an opportunity to predict neurodegeneration in patients with disorders conferring a possible, progressive phenotype. We acknowledge that a limitation of the current assay is small numbers of patients and samples, due to XP rarity. Assay performance could be improved by including other omic modalities, but only if there is an increase in sample size. However, acceptability and value of an early-detection assay can only be realized if it is accompanied by development of prophylactic measures to prevent or substantially delay disease progression. The outlook is potentially optimistic, given observations in this work. Strategies worth considering include reduction of oxidative species and/or maintenance of enhanced UPS activity in neurons, but from a young age in patients that are at risk.

## Online Methods

### Samples

Individuals involved in this study were recruited at the National Xeroderma Pigmentosum (XP) Service at Guy’s and St Thomas’ Hospital and Cambridge and provided formal written consent for the collection and use of their samples in research work. The study was ethically aprouved by Research Ethics Committee in England (REC: Norfolk NRES Committee; REC reference 13/EE/0302) and Scotland (REC: Scotland A Research Ethics Committee; REC reference 14/SS/0096), under the auspices of the Insignia project (IRAS project 116993).

The Blood samples were collected from autosomal recessive XP patients across a broad range of complementation groups, and where possible, from healthy heterozygous relatives. XP patients are followed up longitudinally in the National XP Service and clinical information, including neurological scores are provided in Table S5. All samples were coded and annonimised to protect the donors’ identity.

### Establishing hiPSCs

Human induced Pluripotent Stem Cell (hiPSC) models were generated via the Cellular Generation and Phenotyping facility at the Wellcome Sanger Institute, using the four Yamanaka factors *OCT3/4*, *SOX2*, *KLF4*, *c-MYC* (OSKM). Donors’ peripheral blood mononuclear cells (PBMCs) were first isolated from total blood, the erythroblasts expanded then reprogrammed using a non-integrative CytoTune^TM^ Sendai Reprogramming Kit (Invitrogen^TM^). All hiPSC samples used in the present study were assessed for customary quality control features as previously described^40^, including chromosomal copy number, immunofluorescence markers and expression profiling, to determine pluripotency.

### hiPSC maintenance

All hiPSCs were maintained in feeder-free conditions on Vitronectin FX^TM^ (STEMCELL Technologies) coated plates cultured in Essential E8^TM^ medium (Gibco™). Medium replacement was performed daily, and the cell lines were checked to ensure they were healthy, without signs of spontaneous differentiation. For cell propagation, hiPSCs were treated with 0.5mM UltraPure^TM^ EDTA (Invitrogen^TM^), rinsed with DPBS (Gibco™) and passaged as small cell clumps.

### Neural stem cell (NSC) induction

NSC induction was performed using dual Smad inhibitors (TGF-β Receptor inhibitor and BMP inhibitor) as previously described^40^. The Neural Induction Medium (NIM) consists of: V/V of DMEM/F-12, HEPES (Gibco™) and Neurobasal™ Medium (Gibco™), 1X B-27 Supplement (Gibco™), 1X N2 Supplement (Gibco™), MEM NEAA (Gibco™), 1X Penicillin Streptomycin (Gibco™) in presence of 10µM SB431542 (TOCRIS) and 200nM LDN193189 dihydrochloride (TOCRIS).

hiPSC (one or two replicates for proteomics experiments and independent triplicates for RNA sequencing experiments (Figure S1A)) were dissociated using TrypLE Express Enzyme (Gibco™), and plated on Vitronectin FX^TM^ (STEMCELL Technologies) coated plates at 50,000cells/cm^2^ density in the presence of 1X RevitalCell^TM^ Supplement (Gibco™). hiPSC medium was changed daily until a cell density of 60-75% was reached. The culture medium was switched to NIM medium in presence of 1X RevitaCell^TM^ in the first 24 hours. NIM was changed every day until day 12, marking the end of the neural stem cell induction process.

### Neuronal differentiation

At the end of the neural induction process (day12), the cells were dissociated into single-cell suspension using TrypLE^TM^ Express Enzyme (Gibco™) and plated at high cell density (200,000 cells/cm2) in double-coated plates with Poly-D-Lysine (Gibco™) and 15µg/ml of Cultrex Mouse Laminin I, Pathclear (Bio-Techne). The NIM was switched to Neurons Differentiation Medium (NDM) containing: BrainPhys Neural Medium (STEMCELL Technologies), 1X B-27 Supplement (Gibco™), 1X N2 Supplement (Gibco™), 200nM L Ascorbic Acid (TOCRIS), 20ng/ml BDNF (Cambridge Bioscience), 20ng/ml GDNF (Cambridge Bioscience) with addition of 1X RevitaCell^TM^ for the first 24 hours. The cells were allowed to differentiate for another 30 days (42 days in total). Two-thirds of the medium were changed three times a week.

During the cell differentiation process, cell pellets from all culture replicates were harvested for hiPSC at day 0, differentiating neural stem cell at day 6 and day12, neurons at day27 and day 42 for the RNA sequencing time-series study.

### Immunofluorescence characterization

Immunostaining characterisation was performed at day0 (hiPSC), day12 (NSC) and day27 (neurons) of differentiation to assess differentiation efficiency.

The expression of pluripotency markers of hiPSC was assessed using a commercially available PSC (OCT4, SSEA4) Immunocytochemistry Kit (Invitrogen^TM^). The cells were fixed and stained as recommended by the manufacturer.

NSCs at day12 and neurons at day 27 were stained as follows: the medium was discarded from the plates, and the cells were rinsed gently with 1X DPBS before fixation with a 4% solution of paraformaldehyde for 20 minutes at room temperature (RT). The cells were rinsed twice with 1X DPBS and permeabilised for 20 minutes with 0.1% Triton^TM^ X-100 (Sigma-Aldrich). Non-specific epitopes were blocked with 0.5% BSA solution for 1 hour at RT. Cells were incubated overnight at 4°C with the primary antibodies. For cells at day12, anti-PAX6 and anti Nestin antibodies (Invitrogen™) were used; at day27, cells were incubated with an anti Tubulin beta III (Sigma-Aldrich). The cells were then rinsed three times with 1X DPBS and incubated with the secondary antibody AlexaFluor^TM^ 488 donkey anti-mouse (Invitrogen™). The cells were rinsed three times with DPBS and the nuclei counterstained with NucBlue Fixed Cell Stain ReadyProbes^TM^ (ThermoFisher). The images were acquired within 48 hours using EVOS^TM^ FL Auto 2 microscope (ThermoFisher), and the figures were made using the FigureJ plugin in ImageJ software and final figures panels were made with Addobe Illustrator.

### RNA sequencing

Total RNA was extracted using PureLink RNA Mini Kit (ThermoFisher) according to the manufacturer’s recommendations. cDNA libraries were prepared and sequenced using Illumina Novaseq 6000 technology. We exceeded 20 million read pairs of 150bp paired-end reads for each sample.

### RNAseq analysis

Splice-aware STAR v 2.5.0a^59^ was used to map RNA-seq data to the reference genome. For the human decoy reference genome hs37d5.fa.gz, a genome index was first generated. Then, using the splice junction information from Gencode GTF annotation file v19, fastq files were mapped. The fragments of reads linked with the gene features were then counted using featureCounts v2.0.1^60^. The samples raw counts matrices were then analysed in R version 4.0.4. Differential Gene Expression (DGE) was performed using the DESeq2 R package^61^.

### Quantitative immunofluorescence

NSCs at day 12 derived from NXPD-30, NXPG-32, NXPG-42 and CTRL-33 were seeded in 96 well plates (Perkin) at a cell density of 20 000cell/well and differentiated into neurons for 2 weeks (Day 27) in NDM. The plates were fixed with 4% PFA solution and rinsed three times with 1x, then permeabilised for 10 minutes with Triton 0.2%. The cells were incubated for 1 hour at room temperature with 0.5% BSA solution to block nonspecific epitopes. The following primary antibodies anti-p-PERK (Abcam), anti-GRP78/BiP (Abcam), anti-ATF6 (Abcam), anti-IRE-1α (Abcam), anti-PDI (Abcam), anti-CyclinA (Abcam), anti-PhosphoRB (Abcam), were diluted in 0.5% BSA to desired concentrations and left overnight at 4°C. After washing twice with PBS, secondary antibodies Alexa Fluor® 488 goat anti mouse IgG (Invitrogen), Alexa Fluor™ 488 goat anti rabbit IgG (Invitrogen), Alexa Fluor™ 647 goat anti mouse IgG (Invitrogen), Alexa Fluor® 488 goat anti mouse IgG (Invitrogen) were added for two hours in the dark. Plates were washed twice in PBS, and nuclei were stained with Hoechst (Invitrogen), washed once with PBS, filled with 100μL PBS, and sealed for imaging.

Cells were imaged in the OPERA QEHS (PerkinElmer), with a 60x water immersion objective, on four planes. Maximum automated segmentation was performed using Columbus software (PerkinElmer). Neuronal nuclei were segmented using the Hoechst channel, cells touching the edges were excluded, and the intensity of antibody staining in the nucleus and cell body were measured. All images analysed were created from maximum projections of z-planes. Statistical significance (p. value <0.05) of antibody intensity between samples was assessed using a one way ANOVA with tukey’s multiple comparison test in Prism V.9 (Graphpad). Quantification results were plotted with Prism and further organised into figure panels in Adobe Illustrator.

### XP genotypic correction using CRISPR-Cas12

Undifferentiated hiPSCs of NXPG-32 were maintained in culture as described above until they reached 70-80% confluency. One hour before starting the experiment, the medium was switched to Essential E8^TM^ medium (Gibco™) with RevitalCell^TM^ Supplement (Gibco™) and 2 µM of M3814/Nedisetib (Selleckchem), a potent inhibitor of DNA-dependent protein kinase catalytic subunit (DNA-PKcs). The addition of the later enhances homology-directed repair (HDR) by transiently blocking non-homologous end-joining (NHEJ) activity^62^ leading to a consistent increase in gene editing efficiency. The ribonucleoprotein (RNP) complex was then formed by incubating the Alt-R CRISPR-Cas12 (Cpf1) (IDT) with the crRNA (IDT) for 30 minutes at room temperature (RT) as recommended by the supplier. Vitronectin XF (STEMCELL Technologies) coated 24-well cell culture plate (Sarstedt) was incubated at 32°C for 30 minutes before seeding the CRISPR treated cells.

NXPG-32 hiPSC cells were efficiently dissociated into single cells with TrypLE^TM^ Express Enzyme (Gibco^TM^) and collected in a falcon tube with Essential E8^TM^ medium (Gibco^TM^) and RevitalCell^TM^ Supplement (Gibco^TM^). Cells were counted, and 200,000 cells were centrifuged at 80g for 10 minutes at RT. The supernatant was discarded, and the cells mixed with the RNP complex without introducing air bubbles. A synthetic Alt-R HRD donor oligos template (IDT) was added to the mix, along with the RNP complex that recognises and cuts the targeted DNA sequence. The designed template has a restored wildtype “G” nucleotide that is mutated (deleted) in NXPG-32 and two additional synonymous mutations that modify the targeted DNA sequence (recognised by the crRNA) to prevent the Cas12 enzyme from binding and cutting again after the template was inserted (Figure S2A). The mix (NXPG-32 cells, RNP complex and template) was then transferred in one well of a 16-well nucleocuvette strip and electroporated using SE Cell Line 4D-Nucleofictor X Kit S (Lonza) as recommended. The nucleofection was performed using the 4D-Nucleofector System (Lonza) using the CA137 program. The cells were left at RT for 5 minutes and then plated in one well of the pre-warmed coated 12-well plate in presence of Essential E8^TM^ medium (Gibco^TM^) with RevitalCell^TM^ Supplement (Gibco^TM^) and 2 µM of M3814/Nedisetib (Selleckchem). The cells were then transferred to an incubator at 32°C. 24 hours later, the medium was changed to Essential E8^TM^ medium (Gibco™), and the plate was transferred to an incubator at 37°C. The medium was replaced daily, then 48 hours post nucleofection the cells were treated with 1X of RevitaCell^TM^ for 1 hour, before single-cell dissociation using TrypLE^TM^ Express Enzyme (Gibco^TM^). The cells were centrifuged at 200g for 3 minutes, counted, then 1,000 single cells were seeded in 100mm dish (Corning) coated with Vitronectin XF in Essential E8^TM^ medium (Gibco^TM^) with RevitalCell^TM^ Supplement (Gibco^TM^). The plates were cultured for one week with daily medium replacement. Under a Lynx Stereomicroscope (Vision Engeneering), 25 single colonies with proper round edges were mechanically cut and transferred into 24-well coated plates (Sarstedt). The colonies were cultured until they reached 80% confluency. For each hiPSC clone, half the well was used for cell expansion and the other half was used for DNA extraction. DNA was extracted using Quick-DNA^TM^ Microprep Kit (Zymo Research). The targeted region to edit was PCR amplified and sanger sequenced.

The sanger sequencing data was analysed using SnapGene V6.2 software (SnapGene). Two CRISPR Cas12 edited clones of NXPG-32 were selected to proceed with experiments, NXPG-32-RES, a rescued clone and the NXPG-32-CL2 a clone that was not successfully rescued, although had been through the same process as the rescued clone and would thus serve as the closest control possible. The XPG protein expression was quantified using western blot and proteomics as described further below. Detailed information about the guides and primers is provide in Table S6.

### Western Blot

Proteins were extracted from corrected (NXPG-32-RES) and mutant (NXPG-32-CL2) hiPSC using RIPA buffer (50mM Tris HCl, pH 7.4, 150mM NaCl, 0.1% sodium dodecyl sulfate, 0.5% deoxycholate, 1% NP-40) supplemented with cOmplete^TM^, EDTA-free Protease Inhibitor Cocktail (Roche) and 1% ß-mercaptoethanol. Whole cell extracts were separated by electrophoresis on a gradient polyacrylamide gel. Proteins were transferred onto polyvinylidene difluoride membranes and blocked in 5% skimmed milk dissolved in TBS/0.1% Tween-20. Membranes were incubated with primary antibodies, anti-XPG (Abcam), and anti Actin (Abcam) overnight at 4°C, followed by incubation with Goat Anti-Rabbit IgG H&L (HRP) secondary antibody (Abcam) at room temperature for 1h. Membranes were washed three times in TBS/0.1% Tween-20 after each antibody incubation. Chemiluminescent signal detection was conducted using Pierce^TM^ ECL Western Blotting Substrate as recommended by the supplier (ThermoFisher).

### UVC irradiation and colony formation assay

NXPG-32-CL2 and NXPG-32-RES hiPSC were investigated for functional response to UVC irradiation. Briefly, hiPSC were cultured in two replicate plates until 80-90% confluency as described above. One replicate plate was processed as follows: under the hood, Essential 8 Medium was discarded, and the cells rinsed twice with 1X DPBS. Room temperature DPBS was added to the cells and the plate was transferred to a UVC irradiation chamber. A UVC dose of 0.75 J/m^2^ was applied and the plate was transferred to the hood. The DPBS was replaced with Essential 8 Medium supplemented with Penicillin-Streptomycin. The plate was transferred to the incubator at 37 °C. Both UVC-irradiated and non-irradiated plates were cultured for another 24 hours. Cells were single cell dissociated using the TrypLE Express enzyme, rinsed, counted, and 20,000 cells seeded in 6 well-plates coated with Vitronectin XF. The cells were cultured with Essential 8 Medium supplemented with 1X RevitalCell^TM^ Supplement. The cells were cultured with medium change every day until the NXPG-32-RES cells reached confluency. Under a safety cabinet, the cells were then rinsed with DPBS and covered with 2 ml of a 0.5% crystal violet solution (made with 25% methanol). The cells were incubated at room temperature for 30 minutes and the staining solution was discarded as toxic waste. The cells were rinsed several times with double distilled water. The plates were then inverted and left to dry on an absorbent paper for additional 48 hours. The plates were then imaged. The images were used to calculate the hiPSC growth ratios of UVC-irradiated to non irradiated cells using Image J software and the results plotted in R software.

### Genomic DNA isolation and quantification of modified purine nucleosides

DNA was extracted from cell pellets of neurons cultured for 5 weeks using a high salt extraction method in conjunction with Quick-DNA^TM^ Miniprep Kit (Zymo Research). Cell pellets were resuspended in 200ul of PBS. A nucleic lysis buffer containing 200ul of biofluid cell buffer, 20ul proteinase K, 0.225 mM butylated hydroxytoluene (BHT) and 0.225mM deferoxamine were added to the cells. The lysate was mixed on a thermomixer for 30 minutes at 55°C (850rpm). Equal amounts of genomic binding buffer were added and vortexed. The solution was transferred to a column and centrifuged at 12,000g for 1 minute. The resultant flow through was discarded and a new collection tube added. 450ul of DNA pre-wash buffer was added, centrifuged, the flow-through discarded and a new collection tube added. 800ul of DNA wash buffer was added, centrifuged for 1 minute, the flow-through discarded. 400ul of DNA was buffer was added, centrifuged for 2 minutes and the flow-through discarded. The remaining solution was then transferred to a new labelled eppendorf tube and 50ul of water (pre heated to 55°C) was added. This solution was incubated for 5 minutes and centrifuged for 1.5 minutes at 12,000g. DNA was quantified using a Denovix QFX fluorometer as per manufacturer’s instructions. Ethanol precipitation was performed by adding sodium acetate (3M) at 1/10 the volume of DNA (final concentration 0.3M) and vortexed. The remaining DNA pellet was washed with 2.5x the volume of ice-cold ethanol (100%) to the sample and mixed. The pellet was left at -70°C overnight. The DNA pellet was centrifuged at 12,130 rpm for 30 minutes and the supernatant discarded. The pellet was washed with 1ml 70% ethanol, centrifuged at 12,130 rpm for 15 minutes. The wash and centrifugation steps were repeated, and the supernatant removed. The DNA pellet was dried in a speed vacuum at 55°C for 3 minutes. 10 μg extracted DNA was dissolved in 100 μL of Ar flushed 10 mM Tris-HCl (pH 7.9), containing 10 mM MgCl_2_, 50 mM NaCl, 3 mM deferoxamine, 5 μM ΒΗΤ, 0.2 mM pentostatin and the internal standards were added ([^15^N_5_]–5’*S*–cdA, [^15^N_5_]–5’*R*–cdA, [^15^N_5_]– 5’*S*–cdG, [^15^N_5_]–5’*R*–cdG, [^15^N_5_]–8–oxo–dG and [^15^N_5_]–8–oxo–dA). Benzonase (3 U in 20 mM Tris-HCl pH 8.0, 2 mM MgCl_2_ and 20 mM NaCl), 4 mU phosphodiesterase I, 3 U DNAse I, 2 mU of phosphodiesterase II and 2 U of alkaline phosphatase were added and the mixture was incubated at 37°C. After 21 h, 35 μL of Ar flushed buffer containing 0.3 M AcONa (pH 5.6) and 10 mM ZnCl_2_ were added along with 0.5 U of Nuclease P1 (in 30 mM AcONa pH 5.3, 5 mM ZnCl_2_ and 50 mM NaCl), 4 mU PDE II and 125 mU of DNAse II and the mixture was further incubated at 37 °C for extra 21 h. A step-quenching with 1% formic acid solution (final pH∼7) followed next, the samples were then filtered off by centrifugation through a 3 kDa microspin filter, and were cleaned up and enriched by an Waters Alliance® HPLC–UV system (Waters e2695Separations Module, including a Waters 2998 Photodiode Array coupled with a sample collector) and injected to the LC–MS/MS system loaded with a 2.1 mm × 150 mm Atlantis® dC18, 100 Å column (3 μm particle size, Waters) guarded by a 2.1 mm × 10 mm Guard Column 2pK (Atlantis® dC18 3 μm, Waters). The gradient elution program initiated with 99% of 2 mM ammonium formate (solvent A) and 1% acetonitrile (solvent B) (1 min), increasing solvent B from 1% to 9.8% within 20 min and then immediately to 15% solvent B (5 min), closing with initial conditions for 10 min re-equilibration. The flow rate remained constant at 0.2 mL/min, the injection volume was 50 μL and column temperature was set at 30 °C. Detection was performed in multiple reaction monitoring mode (MRM) using the two most intense and characteristic precursor/product ion transitions for each lesion.

### Mutation accumulation and signature analyses using WGS

Differentiated NSCs (Day 12) were single cell subcloned before whole genome sequencing. For cloning of neural stem cells, the NSCs at P2 were digested using 1x Accutase into single cells and counted with a TC20 Automated Cell Counter (BIO-RAD). Then, 130 cells resuspended in 10 ml NIM medium supplemented with 10 μM Y-27632 (STEMCELL Technologies, 72304) were evenly distributed to a 96-well plate coated with Matrigel at 100 μl/well and cultivated in an incubator at 37 °C with 5% CO_2_. After 24 hours, the 96-well plates were checked under a microscope to record the wells of single cell-derived clones. When the NSC clones grew to 70-100% confluence, they were digested with 1x Accutase and further expanded for approximately 2 months (count beginning from neural induction) before collection for WGS. Four subclones of NXPG-32 and three subclones of CTRL-33 were picked and processed as below.

Genomic DNA from hiPSCs and NSCs were extracted using GenElute^TM^ Mammalian Genomic DNA Miniprep Kit (Sigma‒Aldrich) according to the manufacturer’s instructions. DNA samples passing quality control were sequenced using Illumina X-10 (version 4) technology generating 150bp paired-end short reads equivalent to 120G raw data. The coverage of WGS was ∼30x. Reads for each sample were first mapped to the reference genome of Homo_sapiens (1000Genomes_hs37d5+ensemble_75_trans).

Single base substitutions were called using Cancer Variants Through Expectation Maximization (CaVEMan) algorithm ^63^.The mutations were called by comparing the subclones of NSCs to their parental matched hiPSC. The following criteria were used to exclude low quality substitutions: ASMD > 140 & CLPM = 0.

The 96-channel single substitution catalogues were generated using the “Analyse” tool offered in the online mutational signature analysis website signal (https://signal.mutationalsignatures.com/analyse2) ^64^, and the profiles were plotted in R software. The cosine similarity scores comparing the 96-channels in NXPG-32 and CTRL-33 to SBS18 were calculated using the R package “lsa”.

### Metabolomics sample preparation and analysis

NSCs at day 12 from NXPG-30, NXPG-43 and healthy control CTRL-33 were differentiated into neurons as a monolayer (as described above), 6 well plates were used to generate samples for metabolomics. At the time of metabolite extraction, cells were between 70-90% confluence, and where between 0.4-1 X 10^6^ cells per technical replicate. 500ul extraction buffer per million cells was added and the samples were snap frozen in dry ice then promptly stored at -80 °C until ready for metabolites detection.

Chromatographic separation of metabolites was achieved using a Millipore Sequant ZIC pHILIC analytical column (5 µm, 2.1 × 150 mm) equipped with a 2.1 × 20 mm guard column (both 5 mm particle size) with a binary solvent system. Solvent A was 20 mM ammonium carbonate, 0.05% ammonium hydroxide; Solvent B was acetonitrile. The column oven and autosampler tray were held at 40 °C and 4 °C, respectively. The chromatographic gradient was run at a flow rate of 0.200 mL/min as follows: 0–2 min: 80% B; 2-17 min: linear gradient from 80% B to 20% B; 17-17.1 min: linear gradient from 20% B to 80% B; 17.1-22.5 min: hold at 80% B. Samples were randomized and analysed with LC–MS in a blinded manner with an injection volume was 5 µl. Pooled samples were generated from an equal mixture of all individual samples and analysed interspersed at regular intervals within sample sequence as a quality control.

Metabolites were measured with a Thermo Scientific Q Exactive Hybrid Quadrupole-Orbitrap Mass spectrometer (HRMS) coupled to a Dionex Ultimate 3000 UHPLC. The mass spectrometer was operated in full-scan, polarity-switching mode, with the spray voltage set to +4.5 kV/-3.5 kV, the heated capillary held at 320 °C, and the auxiliary gas heater held at 280°C. The sheath gas flow was set to 55 units, the auxiliary gas flow was set to 15 units, and the sweep gas flow was set to 0 unit. HRMS data acquisition was performed in a range of *m/z* = 70–900, with the resolution set at 70,000, the AGC target at 1 × 10^6^, and the maximum injection time (Max IT) at 120 ms. Metabolite identities were confirmed using two parameters: (1) precursor ion m/z was matched within 5 ppm of theoretical mass predicted by the chemical formula; (2) the retention time of metabolites was within 5% of the retention time of a purified standard run with the same chromatographic method. Chromatogram review and peak area integration were performed using the Thermo Fisher software Tracefinder 5.0 and the peak area for each detected metabolite was normalized against the total ion count (TIC) of that sample to correct any variations introduced from sample handling through instrument analysis. The normalized areas were used as variables for further statistical data analysis using the omu R package^65^.

### Proteomics

Lysis buffer stock solution was made using 1M triethylammonium bicarbonate (TAEB), 5% sodium deoxycholate, 10% isopropanol, 5M NaCl in water. A lysis buffer working solution was made by adding 2ml of the lysis buffer stock solution and 20µl Halt™ Protease and Phosphatase Inhibitor Single-Use Cocktail (ThermoFisher) to an Eppendorf tube. 150µl of the lysis buffer working solution was added to each sample containing a PBS washed cell pellet (approximately 3×10^6^ cells). Samples were then heated at 90°C for 5 minutes and sonicated for a further 5 seconds. Protein concentration was measured with a Bradford assay (Pierce Coomassie Plus) according to the manufacturer’s instructions. Aliquots containing 30ug of total protein in 20µl were reduced with the addition of 2µl of 50mM tris-2-carboxyethyl phosphine (TCEP) and incubated in a heat block at 60°C for one hour. Samples were spun down and then alkylated with 1µl of 200mM freshly prepared iodoacetamide, the solutions were vortexed and spun down. Samples were incubated at room temperature for 30 minutes in the dark. Trypsin digestion was performed by adding 11ul of 100mM TAEB and 6ul Trypsin solution (500ng/µl in 0.1% formic acid), vortexed and spun down. (Protein/trypsin 33:1, trypsin 75ng/µl). Samples were left overnight at room temperature for trypsin digestion. 60ul of 100mM TAEB was added to each sample to bring the volume up to 100µl. Samples were then labelled with TMT pro-16-plex or TMT pro-11-plex reagent vials according to manufacturer’s instructions. 41µl of the TMT reagent was added to each sample and incubated for one hour at room temperature. Hydroxylamine (8ul of 5%) was added to quench the reaction and incubated for 15 minutes. All 16 or 11 samples were combined at equal amounts into one tube. 20µl of formic acid was added and then centrifuged for 5 minutes at 10,000 rpm to precipitate SDC. The remaining supernatant was transferred into a clean tube and dried with a centrifugal vacuum concentrator. “Pooled control” samples were made by creating a “stock pooled control” by combining 50µg of total protein from the control samples made up to 40ul in lysis buffer (11.25ug/µl). 30ug of protein (2.7 µl) of the “stock pooled control” was added to 9 tubes and made up to 20µl with lysis buffer.

### High pH Reversed-Phase Peptide Fractionation and LC-MS Analysis

Peptides were fractionated offline with high-pH Reversed-Phase (RP) chromatography on the XBridge C18 column (2.1 x 150mm, 3.5 μm, Waters) on a Dionex UltiMate 3000 HPLC system. 0.1% (v/v) ammonium hydroxide was used as mobile phase A and 100% acetonitrile, 0.1% (v/v) ammonium hydroxide was used as mobile phase B. The TMT labelled peptides were fractionated with a gradient elution method at 0.2 mL/min, incorporating these steps; for 5 minutes isocratic at 5% B, for 35 min gradient to 35% B, for 5 min gradient to 80% B, isocratic for 5 minutes and re-equilibration to 5% B. Fractions were collected every 42 sec, pooled in 28 fractions, and then vacuum dried. LC-MS analysis was performed on a Dionex UltiMate 3000 UHPLC system coupled with the Orbitrap Lumos Mass Spectrometer (Thermo Scientific). 40 µL 0.1% formic acid was used to reconstitute each peptide fraction and 10 µL was loaded to the Acclaim PepMap 100, 100 μm × 2 cm C18, 5 μm, trapping column at a flow rate of 10 µL/min. Samples were analysed with the EASY-Spray C18 capillary column (75 μm × 50 cm, 2 μm) at 50 °C. Mobile phase A was 0.1% formic acid and mobile phase B was 80% acetonitrile, 0.1% formic acid. The elution method included: for 90 min gradient 5%-38% B, for 10 min up to 95% B, for 5 min isocratic at 95% B, re-equilibration to 5% B in 5 min, for 10 min isocratic at 5% B at a flow rate of 300 nL/min. Precursor ions were selected in the range of 375-1,500 m/z with mass resolution of 120 k, AGC 4×10^5^ and max IT 50 ms with the top speed mode in 3 sec and were isolated for CID fragmentation with quadrupole isolation width 0.7 Th. Collision energy was 35% with AGC 1×10^4^ and max IT 50 ms. Quantification was obtained at the MS3 level with HCD fragmentation of the top 5 most abundant CID fragments isolated with Synchronous Precursor Selection (SPS). Quadrupole isolation width was 0.7 Th, collision energy was 55% and AGC setting 1×10^5^ with 105 ms max IT. The HCD MS3 spectra were acquired for the mass range 100-500 m/z with 50k resolution. Targeted precursors were dynamically excluded for 45 seconds with 7 ppm mass tolerance.

### Database search and protein quantification

Proteome Discoverer 2.4 (Thermo Scientific) using the SequestHT search engine for protein identification and quantification was used to analyse the mass spectra. The precursor and fragment ion mass tolerances were 20 ppm and 0.5 Da respectively. Spectra were searched for fully tryptic peptides with maximum 2 missed-cleavages. Static modifications were designated as TMTpro or TMT6plex at N-terminus/K and Carbamidomethyl at C. Dynamic modifications were designated as Oxidation of M and Deamidation of N/Q. The percolator node was used to estimate peptide confidence and peptides were filtered at q-value <0.01. Peptide confidence was estimated with the percolator node and peptides were filtered at q-value<0.01 based on target-decoy database search. All spectra were searched against reviewed UniProt Homo sapiens protein entries. The reporter ion quantifier node included a TMTpro or TMT-11plex quantification method with an integration window tolerance of 15 ppm at the MS3 level. Only unique peptides were utilised for quantification, using protein groups for peptide uniqueness. Only peptides with average reporter signal-to-noise>3 were used for protein quantification.

### Data analysis using Perseus software

Statistical analysis was performed using Perseus software (version v 2.0.7.0). Samples from each Tandem Mass Tag (TMT) run were normalised to the column sum (by Proteome Discoverer) and log2ratios were computed to the average of the pooled control samples for the corresponding set. Tukey’s biweight was subtracted for each run to account for batch effect. Data from all TMT runs were then combined and average values for the all the control samples combined were subtracted. For cases where there were technical replicates (NXPB-86, XPA-2, XPV-19 and XPE-91) an average value was calculated. Proteins with 1 or 0 valid values were filtered out and samples were normalised by row Z score. Student’s t-test set to a significance value of 0.05 was applied to compare two groups of data. Proteins with a significant difference in abundance between groups were extracted using the threshold of -log p value >1.3 and a student t-test difference of >0.5 or <-0.5 (∼1.4-fold change). Functional classification and pathway analysis were added within Perseus (GOCC, GOBP, KEGG, GSEA). A Fisher’s test with Benjamini-Hochberg FDR threshold of 0.02 was applied to determine enriched pathways. Heatmaps showing proteomics results were generated in R software using normalised s-score scaled data.

### Correlation between quantitative immunofluorescence and proteomics: 1-Regression models

Regression analyses were conducted with the MATLAB (MathWorks) environment using the plsregress function from the machine learning toolbox, Partial least squares regression was selected as the method to help mitigate the influence of co-linearity in the predictor dataset. Model components were selected through 3-fold cross validation. All proteomic expressions were scaled to the total detected mass within each cell line and subsequently scaled across lines prior to model construction. Fit quality was assessed through the r-squared metric.

### 2-Feature Importance to PLSR models

The influence a feature has on a model was estimated through ‘Variable importance to projection’ (VIP) scores calculated as:

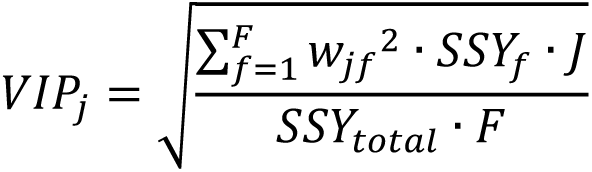

wjf is the weight value for j variable and f component and SSYf is the sum of squares of explained variance for the fth component and J number of X variables. SSYtotal is the total sum of squares explained of the dependent variable, and F is the total number of components. Features with a VIP score greater than 1 were taken as major drivers of the model.

### 3-Peptide expression analysis

Initially peptide expressions were correlated to perk intensity to derive a correlation coefficient, R. ‘Hits’ were defined as those achieving an absolute R > 0.75 corresponding to the critical value for n = 5. Over-representation analysis was conducted on the ‘hit’ set, using the ‘WebGesalt’ server, against all other detected peptides across the gene ontology (biological process), pathway (KEGG) and protein interaction (PPI Biogrid) databases. GSEA used the correlation coefficient as a scoring variable and was conducted across the same databases. Default settings were used in both cases. ‘Hit’ networks revealed through the ontological analysis were visualised in Cytoscape.

### Proteasome Assay

Chromotrypsin-like proteasome activity was measured using the Proteasome Activity Assay Kit (Abcam) as per the manufacturers protocol. Briefly, replicate pellets of 2 million neurons were resuspended in 0.5% NP-40 and centrifuged for 15 minutes at 4°C, 13,000rpm. The supernatants were collected and assayed with and without proteasome inhibitor MG132. Fluorescence was measured at 350/440nm (excitation/emission) using a microplate reader PHERAstar FS (BMG LABTECH). To assess the activity, two readings were made, at 30 minutes, then one hour after incubation with the proteasome substrate. The analysis was performed as described in the manufacturers’ protocol, section “13. CALCULATIONS”. Data was analysed from four independent experiments performed in three to four replicates. Significance between the samples was assessed with a one-way ANOVA (Tukey’s multiple comparison test) using Prism V.9.

### Protein immunoprecipitation using anti-PSMA1

Cells were lysed in 0.5 ml of pre-chilled buffer containing 50 mM Tris pH7.4, 150 mM NaCl and 1% NP-40, supplemented with Halt Protease and Phosphatase Inhibitor Cocktail and Pierce Universal Nuclease. The cells were lysed for 1 h on the rotating wheel at 4C, clarified by centrifugation at 14,000 x g for 30 min at 4C, the supernatant was collected, the protein concentration was determined using the Quick Start Bradford Protein Assay (as per manufacturers protocol) and the protein concentration was adjusted to 0.5 mg/ml. Proteasome immuno-precipitation was performed from 0.5 mg of starting material, using polyclonal rabbit raised anti-PSMA1 antibody coupled to Protein G Dynabeads at the concentration of 50 ug of antibody per 1 mg of protein lysate, incubated for 3 h on the rotating wheel at 4C. Next, flow through was collected and beads were washed in 1 ml PBS 4 times.

The beads were re-suspended in 100 ul of 100 mM triethylammonium bicarbonate (TEAB) and digested with 3 ug of Trypsin at 37C for 18 h, speed vacuum dried. The labelling reaction occurred in 25 ul of 100 mM TEAB and 10 ul TMTpro 18-plex for 1 h according to manufacturer specifications. Samples were combined, speed vacuum dried and fractionated using Pierce High pH Reversed-phase peptide fractionation Kit (according to manufacturer specifications, speed vacuum dried and fractions were resuspended in 0.1% formic acid. LC-MS analysis was performed Dionex UltiMate UHPLC 3000 system coupled with the Orbitrap Lumos Mass Spectrometer (Thermo Scientific). Peptides were loaded to the Acclaim PepMap 100, 100 μm × 2 cm C18, 5 μm, 100 Ȧ trapping column at 10 μL/min flow rate and subjected to a gradient elution on the Acclaim PepMap RSLC (75 μm × 50 cm, 2 μm, 100 Å) C18 capillary column at 45 °C. Mobile phase A was 0.1% formic acid and mobile phase B was 80% acetonitrile, 0.1% formic acid. The separation at flow rate of 300 nL/min was following: 90 min gradient from 5% to 38% B, for 10 min up to 95% B, for 5 min isocratic at 95% B, re equilibration to 10% B in 5 min, for 10 min isocratic at 10% B. Precursors between 375-1,500 m/z were selected with mass resolution of 120 K, Standard AGC and IT 50 ms with the top speed mode in 3 sec and were isolated for HCD fragmentation with quadrupole isolation width 0.7 Th. Collision energy (CE) was 34% with AGC 1×104 and IT 86 ms. HCD MS2 spectra were acquired with fixed first mass of 100 m/z and resolution 50,000. Targeted precursors were dynamically excluded for further isolation and activation for 45 s with 7 ppm mass tolerance.

### Chemical enhancement of UPS activity using PDE4i (Rolipram)

Confluent hiPSC from XP NXPG-32-CL2 and rescued NXPG-32-RES were split into two six well plates coated with Vitronectin XF as described in hiPSC maintenance methods above. One plate (with triplicates of NXPG-32-CL2 and triplicates of NXPG-32-RES) was treated daily with 30 µM Rolipram (Tocris) and one without until cells reached 60-75 % confluency. The hiPSC were then differentiated to neurons as described above.

NXPG-32-CL2 and NXPG-32-RES cells were assessed at multiple differentiation stages Day0 (hiPSC), Day12 (NSC) and Day27 (neurons).

### ROS Quantification Assay

Total ROS levels were measured (in six replicates for each genotype analysed) at different differentiation stages, Day0 (hiPSC), Day12 (NSC) and Day27 (neurons) using ROS-ID Total ROS Detection Kit (ENZO) following the manufacturers’ recommendations. When the cells reached 60-80% confluency the fluorescence was measured using the *Incucyte*® SX5 Live Cell Analysis Instrument (Sartorius). The data was exported, analysed, and plotted using Prism V.9.

### SVM prediction model

Multiple support vector machine (SVM) models were developed to predict neurodegeneration in XP patients using proteomic data. Proteomic data were obtained from neural stem cells that had been differentiated from hiPSCs derived from XP patients with neurodegeneration (eight), without neurodegeneration (five) and with uncertain/undeclared phenotype (due to young age and/or unusual genotype associated with uncertain neurological outcome) (two). Four samples were analysed in duplicate. We explored multiple categories of proteins, including all of the most significantly differentially expressed proteins (DEPs) between patients with and without neurodegeneration, however were concerned about over-fitting, thus explored smaller subsets including the proteins relevant to only the ten most distinguishing biological processes (involving 154 proteins) (Table S5).

The models were implemented in R (version 3.6.2) using svm function from the e1071 library (version 1.7.9). The duplicates were analysed as independent experiments to evaluate the reproducibility of the results. The samples derived from two XP patients with uncertain phenotype were excluded during training. A total of 17 samples, including duplicates, were used for model training. We applied a leave-one-out cross-validation strategy to estimate the distribution of the support vector (SV) coefficients for each selected protein and the probability of each patient to develop neurodegeneration in the derived models. Cross-validation results were also used to identify the optimal number of proteins required to accurately classify the samples.

In each cross-validation iteration, one of the 17 samples was excluded, and two models were derived using the proteomic data of the remaining samples. The first model included the data of the 154 DEPs and the second model used only the data of the proteins with the highest absolute SV coefficients obtained from the first model. Examining the accuracy of the model, if it correctly predicted the classification of the excluded sample, it was set to 1, otherwise it was set to 0. The average accuracy was calculated at the end of cross-validation based on the values defined at each iteration.

In the second model, we performed multiple cross-validation rounds selecting from 1 protein to 16 proteins for each iteration. We selected a maximum of 16 proteins to prevent an overdetermined input matrix. The optimal number of proteins was selected from the cross-validation round with the highest average accuracy. We also evaluated the effect of different seed states (set.seed function) on the average accuracies. We varied the seed states from 1 to 20 and selected the one with highest average accuracy.

The final model was derived using the seed state and the number of proteins at their optimal values. The probability of the XP patients developing neurodegeneration, was predicted using the final model, including the two cases that were excluded because of currently uncertain phenotype.

## Data Availability

All data produced in the present study are available upon reasonable request to the authors

## Acknowledgments

Our heartfelt gratitude goes to XP patients and their families for the generous donation of tissues (blood) to advance research and science. We hope that future work with larger sample cohort can deepen our understanding of neurological implications in XP disease.

## Funding

Wellcome PhD fellowship WT216339/Z/19/Z (to S.M)

Cancer Research UK (CRUK) Advanced Clinician Scientist Award C60100/A23916 (to S.N.Z)

Dr Josef Steiner Cancer Research Award 2019 (to S.N.-Z)

Basser Gray Prime Award 2020 (to S.N.-Z)

CRUK Pioneer Award C60100/A23433 (to S.N.-Z)

CRUK Grand Challenge Award C60100/A25274 (to S.N.-Z)

CRUK Early Detection Project Award C60100/A27815 (to S.N.-Z)

National Institute of Health Research (NIHR) Research Professorship NIHR301627 (to S.N.Z)

NIHR Cambridge Biomedical Research Centre BRC-1215-20014 (to S.N.-Z)

CRUK Centre grant C309/A25144 (to T.I.R, J.C)

Long-term FEBS fellowship (to L.V.-J)

CRUK Programme Foundation award C51061/A27453 (to C.F)

## Author contributions

Conceptualization: CB, SM, SNZ. Data curation: HF, RS, AL, CB, SM. Formal Analysis: CB, SM, SB, TIR, ZK, IJ, JMLD, MGK, AD, CC, SNZ. Funding acquisition: SNZ, JSC, CB, CC, CF. Investigation: CB, SM, GCCK, SB, TIR, ZK, VB, MGK, JY, HC, MY, LVJ, KG, LRK, HF, FR, SZ, CC. Methodology: CB, SM, GCCK, SB, HC. Project administration: SNZ, JSC, CB, CC, CF, CB, SM. Resources: HF, RS, AL, SNZ, CF, CC, CB, JC, CB, SM, YM, JMLD.

Software: CB, IJ, JMLD, YM, AD. Supervision: SNZ. Validation: CB, SM, SB, VB, MGK, JY, TIR, AD. Writing – original draft: SNZ, CB, SM. Writing – review & editing: SNZ, CB, SM, GCCK, SB, TIR, ZK, IJ, JMLD, MGK, JY, HC, MY, FD, LVJ, KG, LRK, AD, HD, RH, CF, CC, RS, AL, CB, JC, HF.

## Competing interests

SN.-Z holds patents on mutational signature-based clinical algorithms not relevant to the research presented in this paper. The other authors declare no competing interests.

## Data and materials availability

Our multi-omics data is not yet deposited in a repository; however, we are happy to provide it as soon as requested by the editors and/or the reviewers. All material relate to this work will be made publicly available before publication.

## Supplementary Materials

## Extended data

Supplementary information. Figs S1 to S5 & Tables S1 to S7

**Fig. S1.**
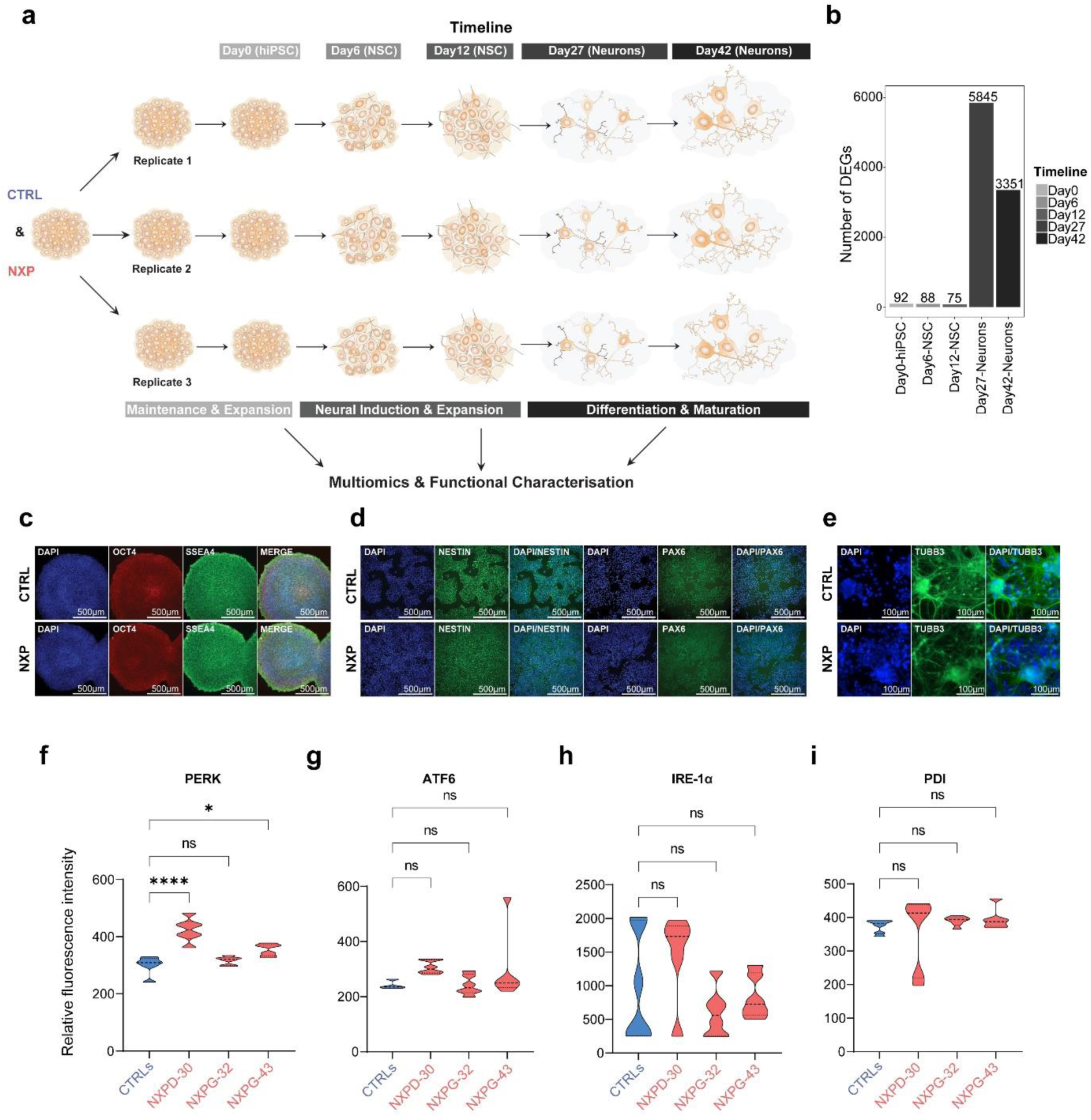
Systematic functional multi-omic study of NXP and controls: experimental design, characterization of various neuronal differentiation stages and validation of upregulation of ER stress/UPR. **(a)** Schematic representation of the experimental design in the present study. **(b)** Number of differentially expressed genes (DEGs) across neuronal differentiation stages. **(c)** Immunofluorescence characterization of human induced pluripotent stem cells (hiPSCs). Scale bards 500µm. **(d)** Immunofluorescence characterization of neural stem cells (NSCs) at day12. Scale bars 500µm. **(e)** Immunofluorescence characterization of neurons at day27. Scale bars 100µm. **(f)** Quantitative immunofluorescence (QIF) of PERK in neurons as day27 (one-way ANOVA, ns= not significant, *: padj<0.05, ****: padj<0.0001). **(g)** QIF of ATF6 in neurons at day 27 (one way ANOVA, ns: not significant). **(h)** QIF of IRE-1α in neurons day 27 (one-way ANOVA, ns= not significant). **(i)** QIF of PDI in neurons at day 27 (one-way ANOVA, ns= not significant).

**Fig. S2.**
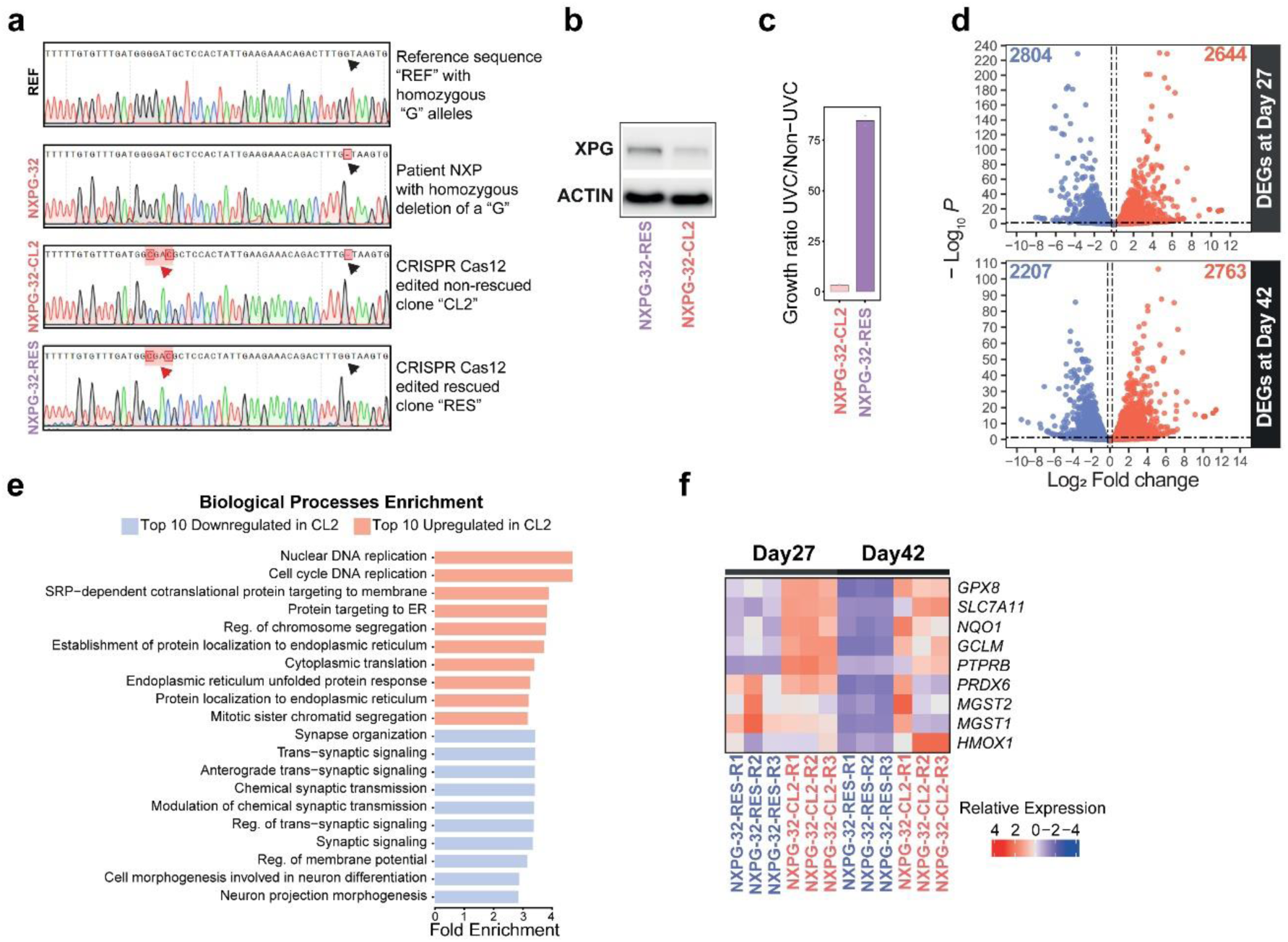
CRISPR-Cas12 complementation of an XPG mutant line reverts the phenotype. **(a)** Sanger sequencing characterization of single-cell subcloned hiPSCs of CRISPR-Cas12 edited XP cells. Black arrows highlighting targeted base; red arrows indicating synonymous mutations also inserted during CRISPR-Cas12 editing, altering the DNA sequence recognized by the crRNA, preventing re-cutting of the edited allele. **(b)** Western blot (WB) characterization of single-cell subcloned hiPSCs of CRISPR-Cas12 edited XP cells. **(c)** Bar plot showing growth ratio of UVC irradiated/non-irradiated CRISPR-Cas12 edited hiPSC. **(d)** Volcano plots showing differentially expressed genes (DEGs) in neurons (day 27 top and day 42 bottom) of an XP case contrasted to complemented cells (padj<0.05). **(e)** Biological Processes GO analysis of the DEGs in XP compared to complemented XP day 42 neurons. **(f)** Heatmap showing difference in expression of NRF2 targets in XP and complemented XP in neurons at day 27 and day 42.

**Fig. S3.**
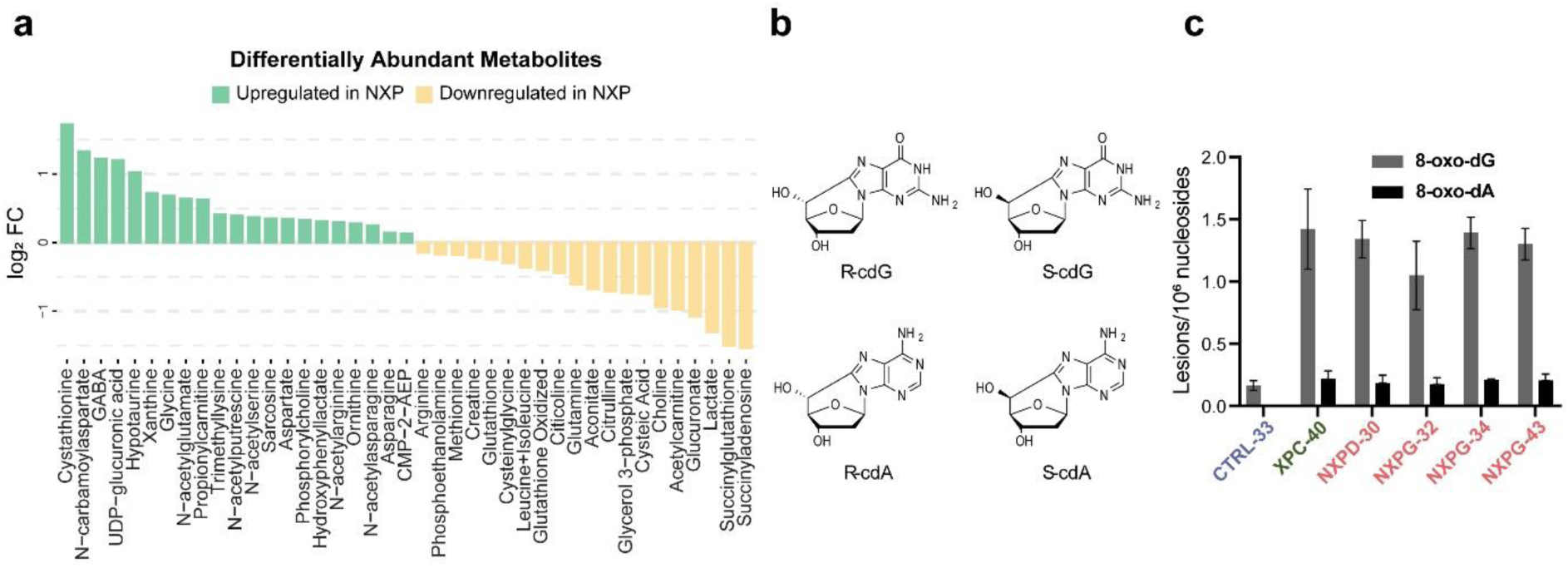
Chronic oxidative stress in XP neurons leads to metabolic adaptation and purine accumulation. **(a)** Differentially abundant metabolites (DAMs) in XP neurons (at day 42) (padj <0.05) compared to controls. **(b)** Structures of 5′,8-cyclo-2′-deoxyguanosine (cdG) and 5′,8-cyclo-2′-deoxyadenosine (cdA) in their 5’*R* and 5’*S* diastereomeric forms. **(c)** Quantification of 8-oxopurine (8-oxo-Pu) in NXP (red), XP without neurodegeneration (green) and controls (blue).

**Fig. S4.**
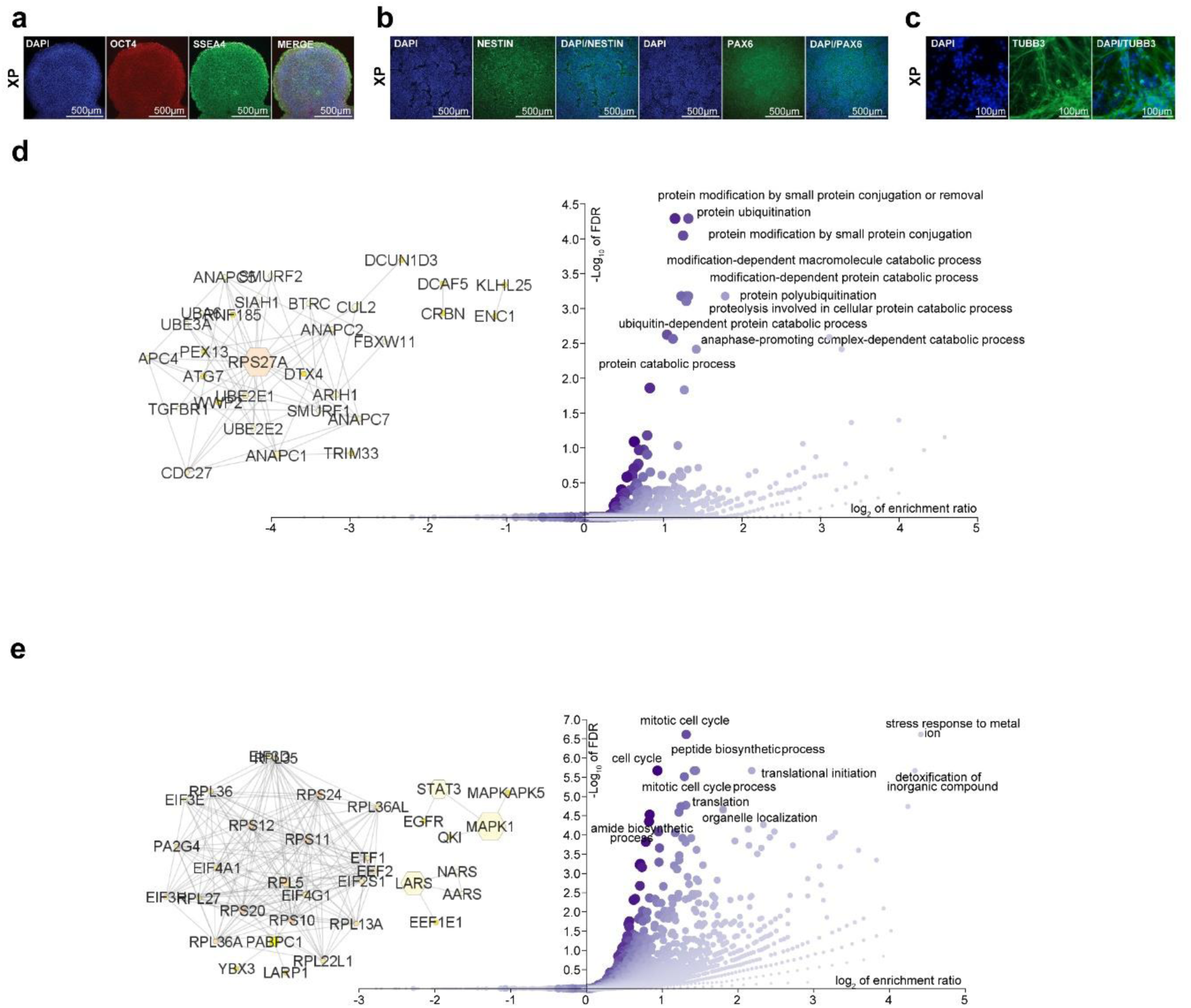
Correlation of proteomics and quantitative immunofluorescence (QIF) links upregulated ER stress with downregulated ubiquitin-related protein catabolism. **(a)** Immunofluorescence characterization of XP-non hiPSCs. Scale bars, 500µm. **(b)** Immunofluorescence characterization of XP-non NSCs at day12 of additional lines made from XP patients and controls. Scale bars, 500µm. **(c)** Immunofluorescence characterization of XP-non neurons at day27. Scale bars, 100µm. **(d)** Over enrichment analysis output showing negatively correlating proteins. Inset shows interaction network of p-PERK (neg) correlates in the ’protein ubiquitination’ theme (GO:0016567). **(e)** Over enrichment analysis output showing themes enriched in strongly positively correlating proteins. Inset shows interaction network of p-PERK correlates in the ’translation’ theme (GO:0006412).

**Fig. S5.**
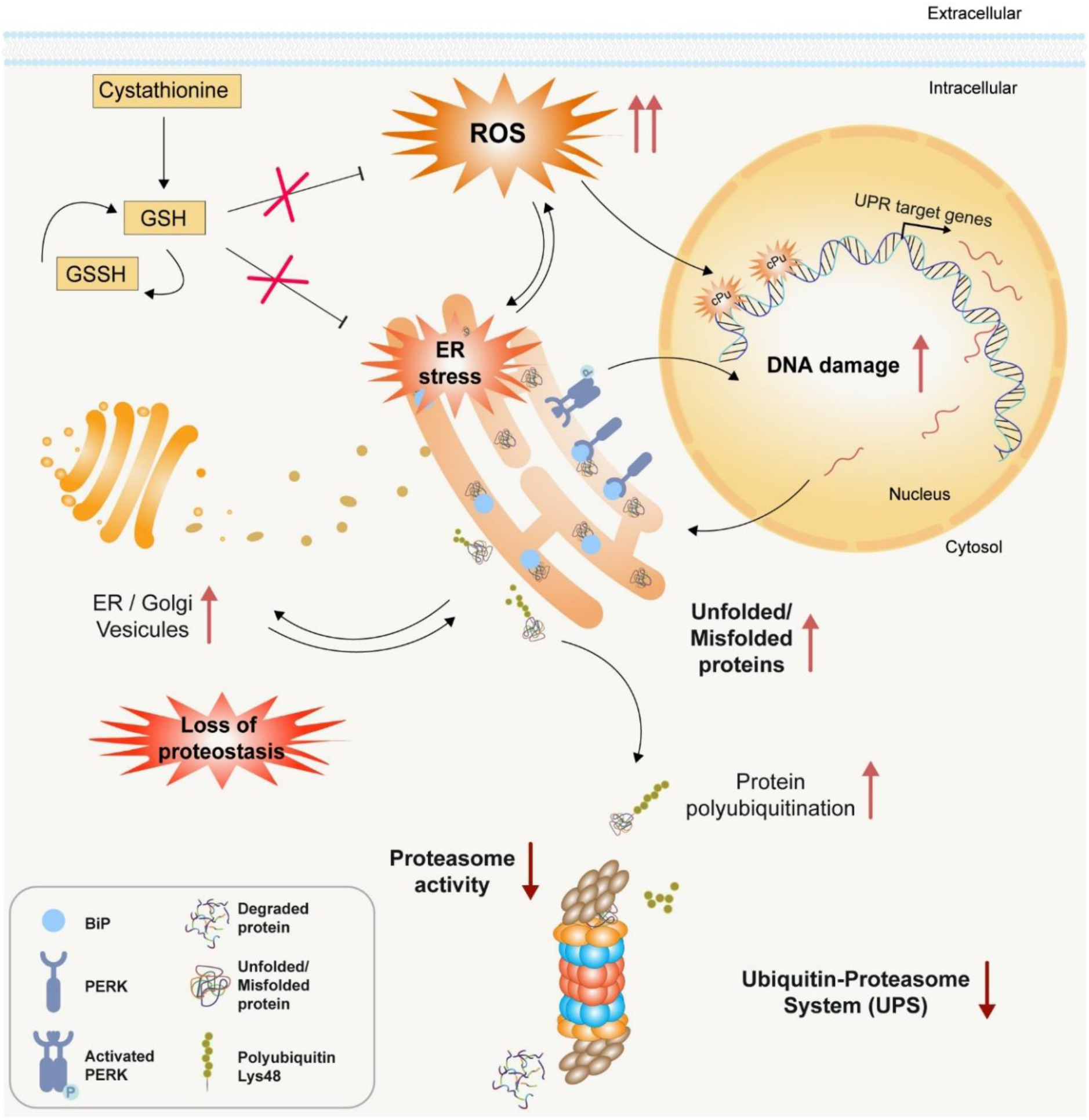
Summary of insights into XP neurodegeneration through functional multi-omics. XP neurons are characterized by ER stress/UPR and loss of proteostasis. In the model of directed neuronal differentiation from hiPSCs of XP patients, evidence of loss of proteostasis is heralded by marked oxidative stress, which remains sustained through differentiation. We find evidence of chronic maladaptation of neuronal metabolism to continuous, substantial oxidative stress, and direct evidence of oxidative DNA damage, with an accumulation of cyclopurine and oxopurine in DNA of XP neurons. While upregulation of ER stress/UPR triggers many appropriate downstream protein clearance pathways including boosting Golgi apparatus activity and markers of de-differentiation, one pathway fails to respond accordingly in patients that do develop neurodegeneration; the UPS. In spite of evidence of appropriate increase in K48 polyubiquitination, we find an inappropriate downregulation of UPS which may be post-transcriptional in origin.

Table S1 to S7 (tables are provided as Datasets of Excel spreadsheets):

Table S1. Bulk RNA-seq data at all differentiation stages, related to Figure 1 and S1

Table S2. Bulk RNA-seq data at Day27 and Day43 for NXPG-32-RES, related to Figure 1I and S2D-F

Table S3. Proteomics data at Day27, related to Figure 3

Table S4. IP anti-PSMA1 data at Day27, related to Figure 4

Table S5. Clinical features of XP-non and NXP, related to Figure 5

Table S6. CRISPR Cas12 rescue, related to Figure 1I and S2D-F

Table S7. List of resources

## Notes

### Competing Interest Statement

SNZ holds patents on mutational signature-based clinical algorithms not relevant to the research presented in this paper. The other authors declare no competing interests.
All other authors declared no competing interest.

### Author Declarations

-Research Ethics Committee in England (REC: Norfolk NRES Committee; REC reference 13/EE/0302)gave ethical approval for this work.

-Scotland A Research Ethics Committee (REC reference 14/SS/0096) gave ethical approval for this work.

